# Human-computer interactions and compassionate healthcare: A Wizard of Oz study using a self-administered AI-assisted cognitive assessment

**DOI:** 10.1101/2024.12.17.24317999

**Authors:** Miini Teng, Martin Yu, Weina Jin, Dae Woo Hwang, David Stitt, Cristina Conati, Giuseppe Carenini, Tal Jarus, Thalia Field

## Abstract

**Introduction:** Artificial intelligence (AI) is increasingly transforming healthcare, however, the interaction between the user, AI, and the user’s environment is poorly understood. To elucidate this interplay and support the delivery of compassionate remote care in Occupational Therapy (OT) practice, we created a self-administered remote AI-powered cognitive assessment, facilitated using the Wizard of Oz method, and measured the experiences of patients, controls, caregivers, and healthcare providers. The Wizard of Oz method uses human-computer interaction (HCI) and user experience (UX) research to simulate the functionality of a system before it is fully developed by creating the illusion of a functioning system by having a human behind the scenes, controlling the system’s responses. Research Questions: 1. How are AI-assisted cognitive assessments experienced by patients, caregivers, healthcare providers, and controls? 2. How can we maintain compassionate care while incorporating technology into healthcare? 3. What are the concerns of healthcare providers using chatbots to administer cognitive assessments?

**Methods:** 6 participants with progressive cognitive decline, 6 healthy controls, 6 caregivers, and 6 healthcare providers were invited to complete a virtual AI-powered cognitive assessment followed by a survey about their experience and other demographical information. Survey questions were separated into 4 scales: Trust, Compassion, Usability, and Care Experience.

**Results:** No statistically significant difference in mean survey scores between participant categories was observed. Factors such as sex, device type, chatbot familiarity, and education had no statistically significant effects. Participants scored statistically significantly lower on the scale Trust (8.09) than on Compassion (8.72). Additionally, those who used the chatbot during the assessment scored statistically significantly lower on the Usability scale compared to those who did not (7.33 vs. 9.20).

**Conclusion:** The findings help to evaluate user experience with virtual AI-based cognitive assessments and provide insights that can inform important design characteristics to improve user experience and compassionate care delivery.

**Author Summary:** As more technological advancements are being achieved in our modern lives today, we continue to see an increasing amount of these technologies in our healthcare system as well. AI is a popular category of these technological advancements and appropriately implementing this powerful tool into day-to-day medicine may prove to benefit our healthcare system. However, we first need to understand our current views on how AI can affect our delivery of care. In this paper, we explored user experience specifically in the context of a cognitive screening tool using wizard of Oz methodology. In brief, our research explores how various stakeholders (patients, control, caregivers, healthcare providers) experience and feel about using a virtual AI-assisted platform for conducting cognitive assessments. We believe that further exploration of AI in medicine and how it can be improved provides an overview of our attitudes towards implementing artificial intelligence into our healthcare system and will also inspire further research for artificial intelligence in medicine.

## Introduction

Mild cognitive impairment is one of the early markers of dementia, Alzheimer’s disease, and other related disorders. Collectively, these disorders are increasing in prevalence and greatly reduce individuals’ functional capacities, having damaging effects on their well-being, and independence, leading to severe societal and economic implications (Prince et al., 2013). This puts extensive strain on patients’ lives, livelihoods, and the healthcare system. The annual cost of dementia to the Canadian economy and healthcare system is over $10 billion (Alzheimer’s Society of Canada, 2016). The detection of mild cognitive impairment can help support timely intervention while patients have largely intact cognitive abilities, better preserving their well-being and independence. Despite the importance of early detection, it is estimated that only 11.4% of individuals who experienced mild cognitive impairment received a timely diagnosis (White et al., 2022) In addition, late detection is especially common in racialized minorities and less resources individuals as many cognitive assessments are biased against less educated and non-English speaking individuals (Sabbagh et al., 2020)

Over the past few years, substantial research has been undertaken to explore the potential of evaluating cognition remotely using telemedicine. This research has been motivated by several factors including a lack of available staff, the COVID-19 pandemic, an aging population, and technological developments. Many already valid and existing cognitive assessments have been adapted for remote administration while others have been developed primarily for this purpose. The use of the Modified Telephone Interview for Cognitive Status (TICS-M) was shown to be sensitive at detecting amnestic mild cognitive impairment (Cook et al., 2009).

Similarly, the Cognitive Telephone Screening Instrument (COGTEL) was shown to be valid and reliable when compared to the Mini-Mental State Examination (MMSE) (Ihle et al., 2017). Other adaptations of existing cognitive assessments such as the MoCA 5-minute protocol have been shown to be equally useful in over-telephone administration (Wong et al. 2015). Likewise, the electronic Self-Administered GeroCognitive Exam (eSAGE), a tablet-based assessment tool, was shown to have a strong association with the paper-based SAGE in detecting mild cognitive impairment and early dementia (Scharre et al., 2017). These studies highlight the important role that remote assessments may play in the future of remote cognitive screening. Remote and self-administered cognitive assessments reduce the burden on healthcare workers and may have the power to create more equitable access for those living in remote or underserviced areas.

Although often more accessible than in-person assessments, many of these remote assessments require administration by a trained professional. Currently, there is a shortage of literature examining patient and healthcare provider experiences with remote assessments. Gaining insight into the obstacles that hinder patient and healthcare provider engagement in remote assessments will contribute to the future development of successful remote cognitive assessments.

A current point of interest in research is focused on utilizing new technologies such as artificial intelligence (AI), machine learning (ML), and virtual reality (VR) in healthcare practice. AI refers to computer systems that behave like a human by engaging in rational problem-solving and decision-making (Russell & Novig, 2020). A systematic scoping review of AI and compassion in healthcare identified 16 articles relating to a theme of human-centered design of AI for healthcare, indicating a significant increase in research in previous years (Morrow et al., 2023). AI has been shown to increase the sensitivity of assessments, allowing for biomarkers of cognitive impairment to be identified. With the use of AI, eye movement data has been shown to be linked to attentional and executive function deficits, indicating it may be useful in increasing the prediction of mild cognitive impairment (Opwonya et al., 2023). Similarly, voice-assist commands to voice-assist systems such as Amazon Alexa were shown to accurately detect various stages of cognitive impairment with AI analysis (Kurtz et al., 2023). These studies highlight the potential for the development of repeatable and remote AI-based methods of cognitive assessment. Additionally, some AI-based assessments have been designed to be language-independent which reduces biases associated with non-native speakers and individuals with expressive aphasia. The Integrated Cognitive Assessment (ICA) is both self-administered and language-independent. Using AI, the ICA showed no learning bias and was shorter in duration than the Brief International Cognitive Assessment for MS (BICAMS) (Khaligh-Razavi, et al., 2020). AI has the potential to increase the sensitivity and speed of cognitive assessments while reducing cost and language barriers.

ML is a branch of AI that focuses on the use of datasets to improve decision-making capabilities through a process akin to human learning. ML and the use of large datasets may further increase the screening capabilities of future AI-powered assessments (Battista et al., 2020). Deep learning programs such as Xception have been shown to differentiate the faces of patients with mild dementia from those without dementia, indicating the possibility of developing facial biomarkers for dementia (Umeda-Kameyama et al., 2021). VR technology has been used in assessments to create a more immersive and functional assessment in spaces where limited resources are available. VR was shown to be a feasible medium for assessing cognition and detecting mild cognitive impairment (Zygouris et al., 2017). Presently, much of the literature on utilizing these technologies in healthcare is focused on the development of biomarkers and new assessments. There is limited research investigating user and healthcare provider experiences and perceptions with AI-based assessments. A scoping review of perceptions of AI in healthcare to increase adoption identified a lack of studies on the perceptions of AI and a shortage of healthcare professionals to report on the attitudes of the public (Chew & Achananuparp, 2022). Further understanding of user experience will help support designs that are most likely to be adopted by users and healthcare providers, which will in turn foster usability, garner trust in results, and ensure compassionate care. A systematic review of AI-powered chatbots in health promotion identified mixed results with respect to acceptability and usability, with low satisfaction rates (Aggarwal et al., 2023). Some issues identified in the review were poor fluidity of conversations, a lack of technological skills of participants, technical challenges, and an absence of emotional connection. While AI-powered assessments and interventions have gathered much interest, deploying these sorts of technological solutions requires an understanding of the interactions between the people involved, the AI-powered tasks, and the environments in which these tasks take place.

Significant contemporary research has been conducted on the user experience with remote, non-AI-based, self-administered cognitive assessments. One of the main challenges in self-administered and remote assessment is that patients must be able to understand and operate the equipment required for the assessment. Recent research topics have centered around the creation of user interfaces that are simple and user-friendly. Utilizing the Systems Usability Scale (SUS) to inform design, a robot-administered, AI-based test inspired by the MoCA was created and shown to be positive with respect to usability for participants in terms of its consistency, ease of use, and confidence of use among other metrics (Di Nuovo et al., 2019a) The SUS helps to highlight important design implications when developing an assessment in which participants self-administer. Further studies from this group found that the robot-administered, AI-based, MoCA-inspired assessment gave significantly lower scores than corresponding MoCA scores due to rigid administration procedures and improper speech recognition (Di Nuovo et al., 20199b). Human-robot interactions that were found to affect administration were crashes of the interface, unclear pronunciation of instructions, too little time to complete tasks, and improper placement of the sheet in front of sensors. These issues highlight the many human-computer interaction challenges that affect the validity of remote and self-administered cognitive assessments. It is crucial to understand the demographics, computer literacy, and resources such as the internet and available devices of a target population when designing an assessment. Data on demographics of telehealth users indicated that areas with broadband internet had decreased use of smartphone-accessed virtual care (Broffman et al., 2023). Additionally, it was found that older men and people of marginalized communities were likely to access virtual care via smartphone due to a lack of other devices. This indicates that impactfully reaching more remote communities may require a more flexible assessment design that can be run equally effectively on different devices. This research highlights the complex interplay between the users, their attitudes, and the assessment tool in determining their experience.

The Wizard of Oz study design has been utilized in the design phase of healthcare chatbots to discover the common questions asked, determine how the chatbot should engage the participants, and collect real-time information on participant interactions. (Minian et al., 2023) The Wizard of Oz is a widely accepted method for designing and appraising systems that require human-computer interaction, such as chatbots. (Hanington & Martin, 2019) A scoping review of chatbot development in healthcare recommended the use of the Wizard of Oz method when developing new projects. (Sadasivan et al., 2023) By exploring the role of human-computer interactions on user experience we can inform the design of improved remote, AI-based cognitive assessments. Presently, there has been a dearth of research relating to human-computer interactions and user experience with respect to self-administered, remote, AI-based cognitive assessments. To help address this gap in research, this study applied the Wizard of Oz methodology to understand the interplay between demographic factors and user experience when participating in a virtual iteration of an existing AI-powered cognitive assessment called CANARY.

CANARY is an acronym that stands for clinical data, natural language processing and eye tracking for dementia risk stratification. The CANARY assessment was designed by an interdisciplinary team of neurologists and computer scientists at the University of British Columbia (UBC). It aims to use AI and ML to detect mild cognitive impairment and early-stage neurodegeneration using eye tracking and speech data during a series of simple tasks such as describing an illustrated scene, reading a paragraph, and recalling a memory. The original CANARY study is administered in person at the clinic with the assistance of a staff member to support the setup and administration. The iteration designed for this study contains the same tasks but is virtual and self-administered and AI chatbot-assisted. This study aimed to understand how an AI-powered cognitive assessment is experienced by users, including healthcare providers and caregivers. An additional aim was to assess how compassionate care can be supported while incorporating technology into healthcare. The final aim was to identify concerns and opportunities of healthcare providers using AI-powered cognitive assessments. The findings aim to better understand the interplay between the patient, AI, and the user environment with the aim of supporting the delivery of compassionate remote care.

## Methodology

### Study Design

This study aimed to evaluate the human-computer interactions involved in the remote and self-administered iteration of a previously developed AI-powered cognitive screen (CANARY). An additional aim was to explore user experience and perception for each participant group; patient (mild cognitive impairment), caregiver, healthcare provider, and control. Our study involved 6 participants in each of these participant groups. Human-computer interactions were assessed using survey data and naturalistic observations involving observing subjects in their natural environment. Upon completion of the assessment, participants filled out a survey about their experiences during the assessment as well as demographic and environment-related questions such as age, sex, device type, familiarity with AI chatbots, and level of education.

Additionally, caregivers were asked to self-report the number of years in a caregiving role and healthcare providers were asked to self-report the number of cognitive assessments administered. This data was analyzed using SPSS version 29.0.1.0 to better understand the role of the participants’ demographics and environment in remote and virtual assessment experiences.

### Participants

Participants were enrolled through a sample of convenience, using email mailing lists compiled from previous studies who consented to enroll in future study mailing lists (including the CANARY study), emails to different healthcare and non-profit organizations, word of mouth, posters, and recruitment at local memory clinics and other on-site locations. Ethical approval was obtained through the UBC Behavioural Research Ethics Board. Additional approval to the Vancouver Coastal Health Research Institute (VCHRI) was obtained for the purpose of contacting Vancouver Coastal Health employees via email, placing study posters in Vancouver Coastal Health spaces, and recruiting on Vancouver Coastal Health property. Interested individuals were asked to email a study-specific email. Those who emailed about their interest were followed up by study team members at UBC via their UBC-affiliated emails. Participants were sent a consent form that outlined information about the study purpose, study procedures, risks and benefits, confidentiality, and compensation. Consent forms were obtained through electronic copies. Once the consent forms were received, participants were sent a Calendly link to book an assessment time slot with a study team member. The inclusion and exclusion criteria for each participant group are listed below.

### Inclusion Criteria

#### Patients

1. 19 years or older
2. Either:

a. At high risk of progressive neurodegenerative disease or progressive cognitive decline (e.g. gene positive for familial early onset Alzheimer’s disease, gene positive for Huntington’s disease, mild cognitive impairment, subjective memory complaints, Rapid Eye Movement sleep behavior disorder (RBD) or otherwise), or
b. manifest neurodegenerative disease (e.g. clinical diagnosis of Alzheimer’s Disease, mild cognitive impairment (MMSE score 19-25), Huntington’s disease, frontotemporal dementia, Parkinson’s disease, or otherwise)
3. Fluent in English
4. Ability to provide informed consent
5. Patient or is able to carry on a spontaneous conversation (neurodegenerative disease is not so advanced that they are unresponsive)
6. Able to attend study assessments

#### Healthy controls

1. 19 years of age or older
2. Fluent in English
3. Ability to provide informed consent
4. Able to attend study assessments, outlined below

#### Caregivers

1. 19 years of age or older
2. Providing care for a patient who can participate in the study (part of the inclusion criteria above)
3. Fluent in English
4. Can provide informed consent for themselves
5. Can attend and participate in the cognitive assessments

#### Healthcare Professionals

1. All clinicians and allied healthcare professionals (ex. Neurologists, Occupational Therapists, Psychiatrists, Nurses, etc.) who are involved in cognitive screening
2. Currently practicing in their respective fields

### Exclusion Criteria

#### Patients

1. Severe cognitive impairment (MoCA score of <10)
2. Dysphasia secondary to causes other than dementia/MCI
3. Currently diagnosed with a psychiatric condition that is undergoing active psychiatric follow-up with changes in treatment occurring within the last 18 months
4. Severe eye or vision condition that would affect accuracy of eye-tracking.5. 5. Not fluent in English.

#### Healthy Controls

1. Previous brain injury, stroke, TIA or other primary neurological condition (excluding migraine)
2. Currently diagnosed with a psychiatric condition that is undergoing active psychiatric follow-up with changes in treatment occurring within the last 18 months
3. Severe eye or vision related condition that would affect accuracy of eye-tracking.
4. Not fluent in English.

#### Caregivers

1. Not fluent in English.
2. Cannot provide informed consent for themselves or patients receiving care.

#### Healthcare Professionals

1. Not currently practicing (ex. retired, leave of absence)
2. Healthcare professionals who’s routine work does not involve cognitive assessments.

## Procedures

### Assessment Design

This iteration of the CANARY experiment was hosted on a private website, password protected, and hosted on a Canadian server. The assessment was partially automated using the SpeechSynthesisUtterance text-to-speech software and the Web Speech API speech-to-text software developed by Mozilla. This software allowed written prompts on the screen to be read aloud, requiring participants to click onward when they had finished each task. The tasks performed are identical to the original CANARY experiment and consisted of 4 short tasks performed over 10 minutes. The tasks performed by participants were ordered as follows: 1. fix eyes on a cross for 10 seconds, 2. describe an illustrated scene aloud, 3. read a paragraph aloud, and 4. describe a positive life event aloud. With respect to the Wizard of Oz, if participants wished to ask a question to the chatbot during the assessment, they would click a question mark icon on the assessment screen, then click a microphone icon and ask their question aloud or by typing. This question was transcribed to text and an answer was generated for the wizard (a study team member) by an AI chatbot. This AI chatbot was powered by the GPT-3 extension created by OpenAI and trained with the CANARY assessment using few-shot training. The participant question and AI-chatbot answer were displayed for the wizard. The wizard would discard the AI-generated answer and type an appropriate answer to the participant’s question. The wizard-generated response would then be displayed in text on the participant’s assessment screen and read aloud. This procedure would occur without alerting the participant to the fact that answers were produced by study team members. Wizards had a list of commonly asked questions and appropriate answers to support their ability to answer quickly. The Wizard of Oz design was incorporated to understand what questions participants asked, what answers an AI-chatbot would provide, illuminate the timing and context of questions to better understand barriers in the user experience, and ensure appropriate answers were provided to the participants.

### Study Protocol

Participation took place during June and July 2023 with participants meeting with a study team member over Zoom during their scheduled assessment time (30 minutes). Participants performed the virtual and self-administered iteration of the CANARY experiment with their own devices (laptop, tablet, desktop, etc.) from their location of choice, most times in their homes.

Study team members introduced themselves, briefly reiterated the purpose of the study, and described the study procedure. Participants were also requested to have their webcams on and to share their screens and audio during the assessment for recording purposes. Once this was set up, study team members informed participants that they may begin the assessment and that any questions they may have during the assessment should be directed to the chatbot within the assessment to simulate a fully virtual assessment. The chatbot within the assessment introduced itself as Al and could be accessed by clicking a question mark icon on the screen. At the beginning of the assessment, participants are reminded to remain seated, keep their hands at their sides, and keep their eyes on the screen for the duration of the assessment. Upon completion of the assessment, participants filled out a Qualtrics survey with questions about their age, sex, device type, familiarity with AI chatbots, and level of education. Additional survey questions focused on user experience and were divided into 4 main categories, or scales: Trust, Compassion, Usability, and Care Experience. The survey questions about user experience were inspired by the Schwartz Center Compassionate Care Scale, Systems Usability Scale, Hospital Consumer Assessment of Healthcare Providers and Systems, Short-form Hong Kong Inpatient Experience Questionnaire, Patient Experience Questionnaire, and the Technology Acceptance Toolkit.

All electronic files of study documents, Zoom recordings of assessments, and survey responses were stored securely in separate password-protected folders on UBC OneDrive, which is encrypted. The folders in UBC OneDrive will only be accessible through each researcher’s UBC email account. No data was saved on the researchers’ personal laptops. All raw data was only available to team members. All participant identifiers were removed to protect the participants’ confidentiality and anonymity. After all data was collected and the results were finalized, the study participants were contacted and informed that they were not in fact interacting with an AI system, but the responses were monitored and adapted by study team members during the assessment.

## Measures

Survey responses about the age, sex, device type, familiarity with AI chatbots, level of education, home environment, and caregiving/cognitive assessment experience were measured on a nominal scale through drop-down menus with prompts and options for typed responses if prompts were not applicable. Responses for participant experience were recorded on an ordinal scale from 1-10 (strongly disagree – strongly agree) with an option for not applicable.

The 17 survey questions on user experience were separated into 4 main scales as shown below.

3 questions related to the theme of ‘Trust’.

1. I would trust the validity of this assessment.
2. I am suspicious of the assessment.
3. My privacy was respected during the assessment.

3 Questions related to the theme of ‘Compassion’.

1. I felt listened to during the assessment.
2. I think this assessment treats someone as a person not as a disease.
3. I felt respected during the assessment.

5 Questions related to theme of ‘Usability’.

1. The information was conveyed in a way that is understandable.
2. It was difficult to ask questions during the assessment.
3. The communication was done in a timely and sensitive manner.
4. I consider the assessment easy to use.
5. Overall, I had a very positive experience with this assessment.

6 Questions related to the theme of ‘Care experience’.

1. During this assessment, I was able to get help when I needed it.
2. I can see this assessment being used in healthcare.
3. I feel that the assessment is secure and safe.
4. I feel the assessment took a reasonable amount of time.
5. I would be likely to recommend this assessment to my friends, family or colleagues.
6. Using this assessment would be a helpful tool in managing my health.

Scales or “Themes” were analyzed as a unit to understand the patient experience in that domain of the user experience.

## Data Analysis

Data were analyzed and displayed visually using Microsoft Excel and Statistical Package for the Social Sciences (SPSS) version 29.0.1.0. Survey data from Qualtrics was exported in the form of an SPSS file. Overall survey scores as well as mean values for each scale; Trust, Compassion, Usability, and Care Experience were analyzed for statistical significance between and within the 4 participant groups: patients, healthy controls, caregivers, and healthcare providers. These scores were then also analyzed with regards to demographic information such as sex and level of education as well as environmental information such as device type and familiarity with chatbots. Credibility was supported through triangulation; participants were invited to participate in focus groups to discuss their experience with the assessments. Focus group analysis was not performed prior to the writing of this manuscript and thus the results are not displayed. However, focus group participation will increase the credibility of the representation of the user experience by engaging with participants in the different context of a free-flowing, guided discussion.

Member checks will be performed during focus groups to ensure the accuracy of the data obtained. Peer review from project facilitators was also used to support the congruence between the raw data obtained and the findings of the final report.

## Results

### Demographics

Overall, 24 Participants (14 females and 10 males) took part in the study (Figure 1). There were 6 participants in each of the 4 participant categories (patient, caregiver, healthcare provider, and healthy control). The age range for participants was between 28 and 79 years of age (Table 1). The mean age of each participant group was as follows: patients – 65.83 years old, caregivers −68.00 years old, healthcare providers – 39.50 years old, and controls - 69.67 years old (Table 2).

**Figure 1.**
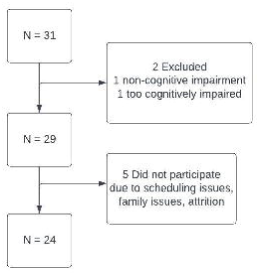
Participant inclusion flowchart.

**Table 1.**
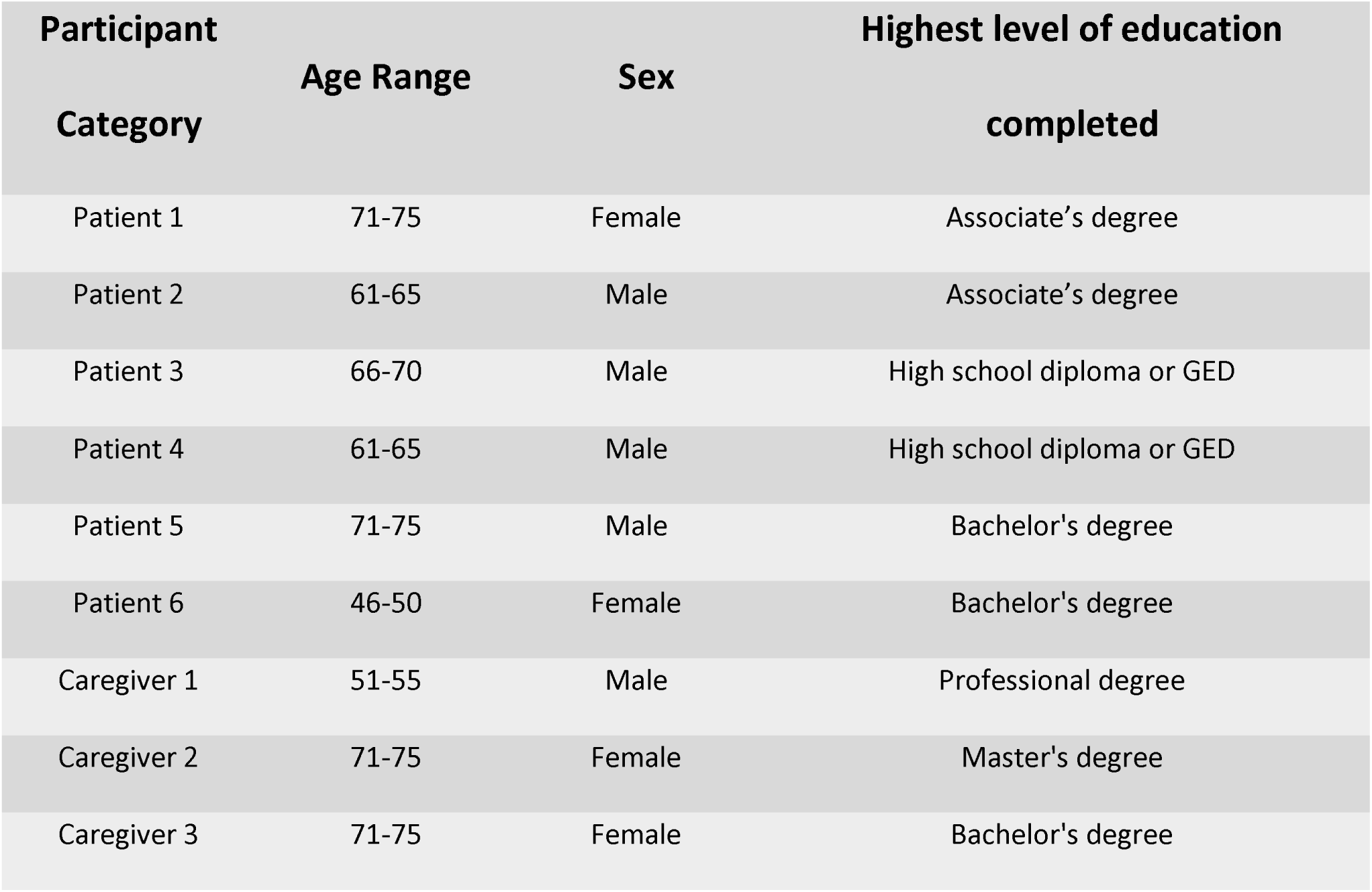
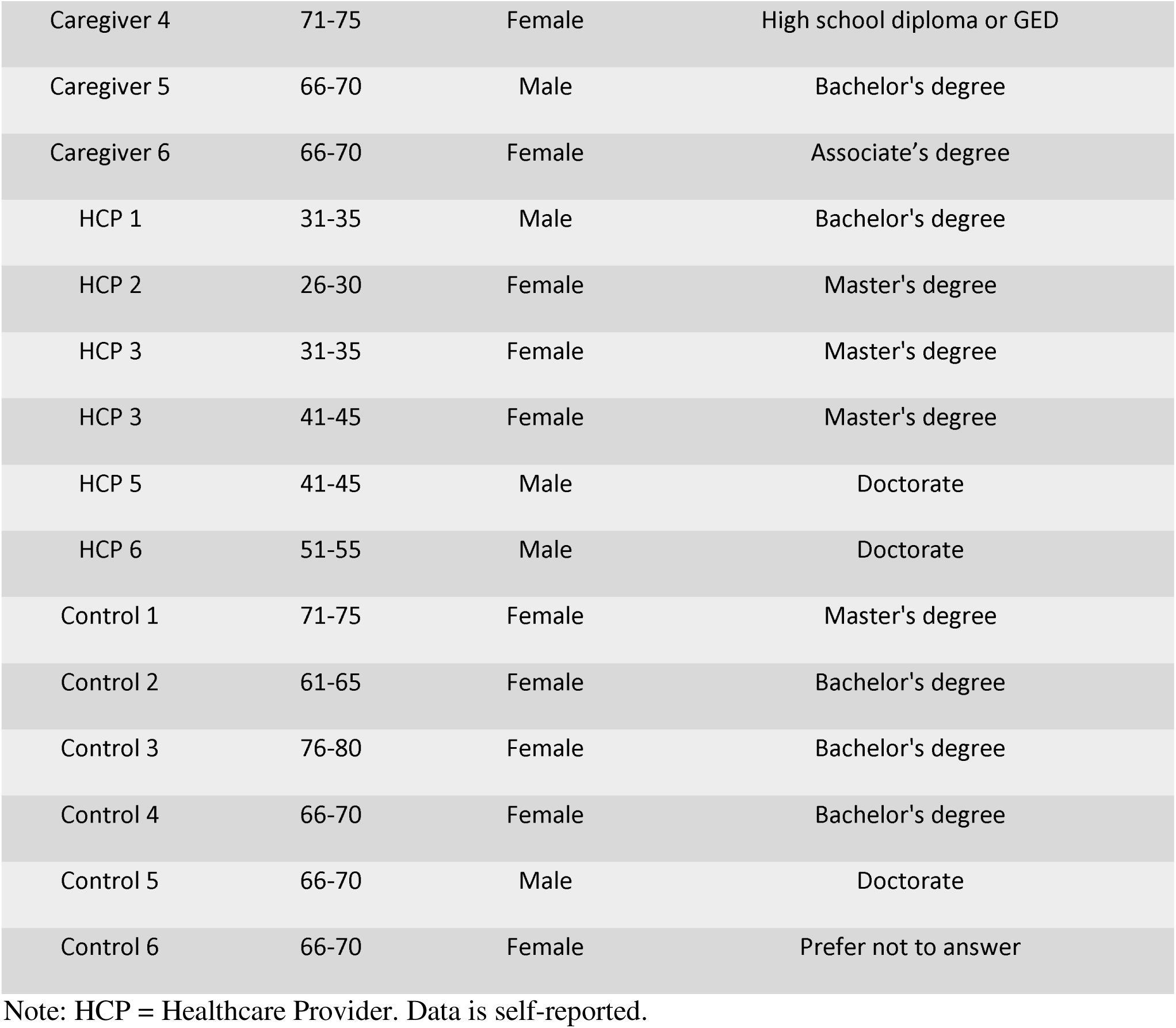
Participant Demographics.

| Participant Category | Age Range | Sex | Highest level of education completed |
| --- | --- | --- | --- |
| Patient 1 | 71-75 | Female | Associate's degree |
| Patient 2 | 61-65 | Male | Associate's degree |
| Patient 3 | 66-70 | Male | High school diploma or GED |
| Patient 4 | 61-65 | Male | High school diploma or GED |
| Patient 5 | 71-75 | Male | Bachelor's degree |
| Patient 6 | 46-50 | Female | Bachelor's degree |
| Caregiver 1 | 51-55 | Male | Professional degree |
| Caregiver 2 | 71-75 | Female | Master's degree |
| Caregiver 3 | 71-75 | Female | Bachelor's degree |
| Caregiver 4 | 71-75 | Female | High school diploma or GED |
| Caregiver 5 | 66-70 | Male | Bachelor's degree |
| Caregiver 6 | 66-70 | Female | Associate's degree |
| HCP 1 | 31-35 | Male | Bachelor's degree |
| HCP 2 | 26-30 | Female | Master's degree |
| HCP 3 | 31-35 | Female | Master's degree |
| HCP 3 | 41-45 | Female | Master's degree |
| HCP 5 | 41-45 | Male | Doctorate |
| HCP 6 | 51-55 | Male | Doctorate |
| Control 1 | 71-75 | Female | Master's degree |
| Control 2 | 61-65 | Female | Bachelor's degree |
| Control 3 | 76-80 | Female | Bachelor's degree |
| Control 4 | 66-70 | Female | Bachelor's degree |
| Control 5 | 66-70 | Male | Doctorate |
| Control 6 | 66-70 | Female | Prefer not to answer |

Note: HCP = Healthcare Provider. Data is self-reported.

**Table 2.** Mean Age by participant category.

| Age by participant category (Mean ± Standard Deviation, Range) |  |  |  |
| --- | --- | --- | --- |
| Patient | Caregiver | Healthcare Provider | Control |
| 65.83 ± 9.15, 25 | 68.00 ± 7.40, 20 | 39.5 ± 8.34, 23 | 69.67 ± 6.50, 18 |

## SPSS Analysis

### Effects of age

Age was not quantitively analyzed through ANOVA because the age distribution between participant categories was wide. More specifically, while the mean age for patients, caregivers, and controls were in the 60’s, the mean age for healthcare providers was 39.5 (Table 2). Therefore, age would be a confounding variable when comparing age to overall survey scores and analyzing age against survey scores would not accurately represent an age-related difference of scores.

### Effects of scale

The results of the repeated measure ANOVA indicated that there was no overall statistically significant differences of mean scores between the 4 scales (Trust, Compassion, Usability, Care Experience) with medium effect size (Table 3). That is, when comparing the overall scores of the 4 scales to one another, no one scale scored statistically higher or lower than the other scales (Figure 2). However, when comparing the scales individually, the results show that there is a statistically significant difference between the mean scores of scale 1 (Trust) and scale 2 (Compassion) with large effect size. Participants scored significantly higher on questions related to compassion compared to questions related to trust on the survey (Figure 2). No other statistically significant differences were found between any other individual scales.

**Figure 2.**
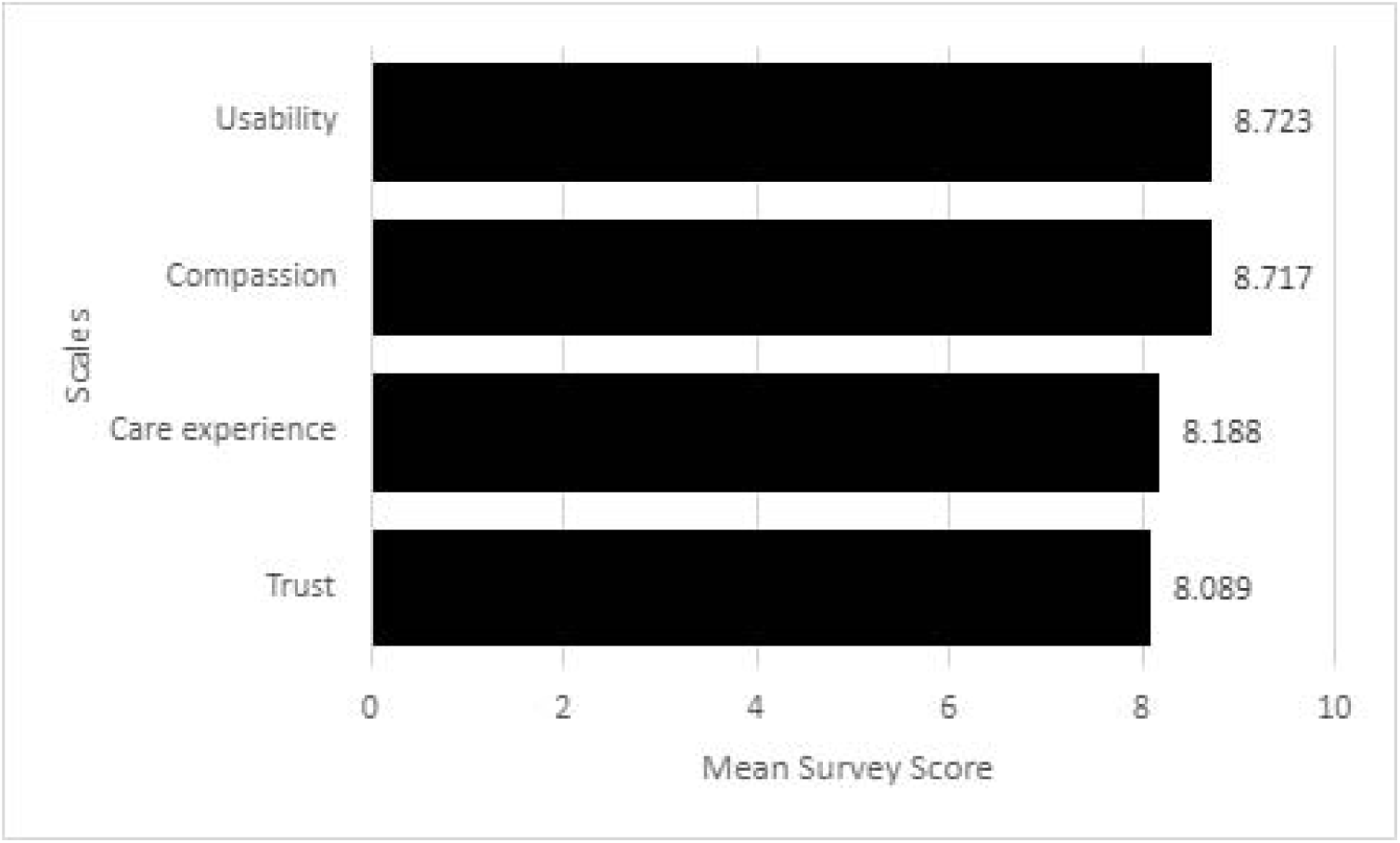
Overall mean survey scores across the 4 scales.

**Table 3.**
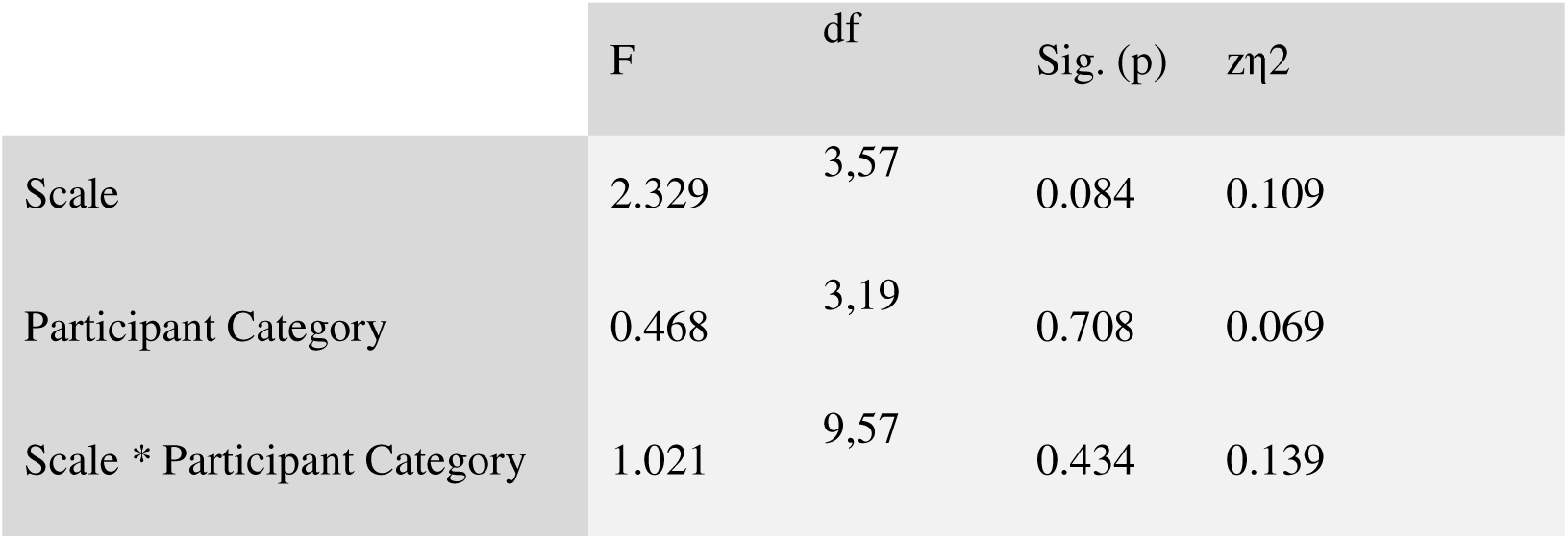
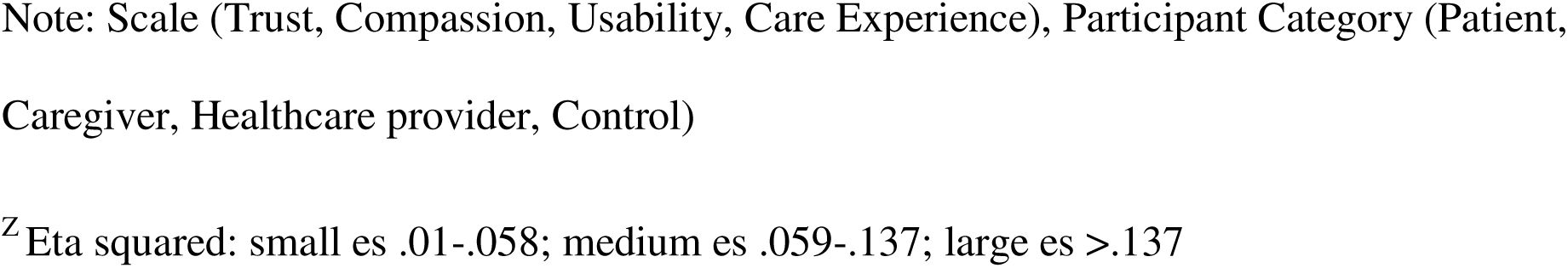
Results of the ANOVA of Scale by Participant Category.

*Results of the ANOVA of Scale by Participant Category*
| | F | df | Sig. (p) | $\eta^2$ |
| --- | --- | --- | --- | --- |
| Scale | 2.329 | 3,57 | 0.084 | 0.109 |
| Participant Category | 0.468 | 3,19 | 0.708 | 0.069 |
| Scale * Participant Category | 1.021 | 9,57 | 0.434 | 0.139 |

Note: Scale (Trust, Compassion, Usability, Care Experience), Participant Category (Patient, Caregiver, Healthcare provider, Control)
<sup>z</sup>Eta squared: small es .01-.058; medium es .059-.137; large es >.137

### Effects of participant category

The results of the repeated measure ANOVA indicated that there was no overall statistically significant difference of mean scores on the survey between the 4 participant categories (patient, caregiver, healthcare provider, control), with medium effect size (Table 3). That is, all 4 participant categories scored similarly when looking at the overall mean scores on the survey (Figure 3). When comparing the scales individually within each participant group, the results show that there is no statistically significant difference on the mean survey scores of each scale. That is, within each participant group, participants scored similarly across all 4 scales (Figure 4).

**Figure 3.**
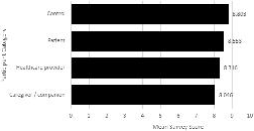
Overall mean survey scores across participant categories.

**Figure 4.**
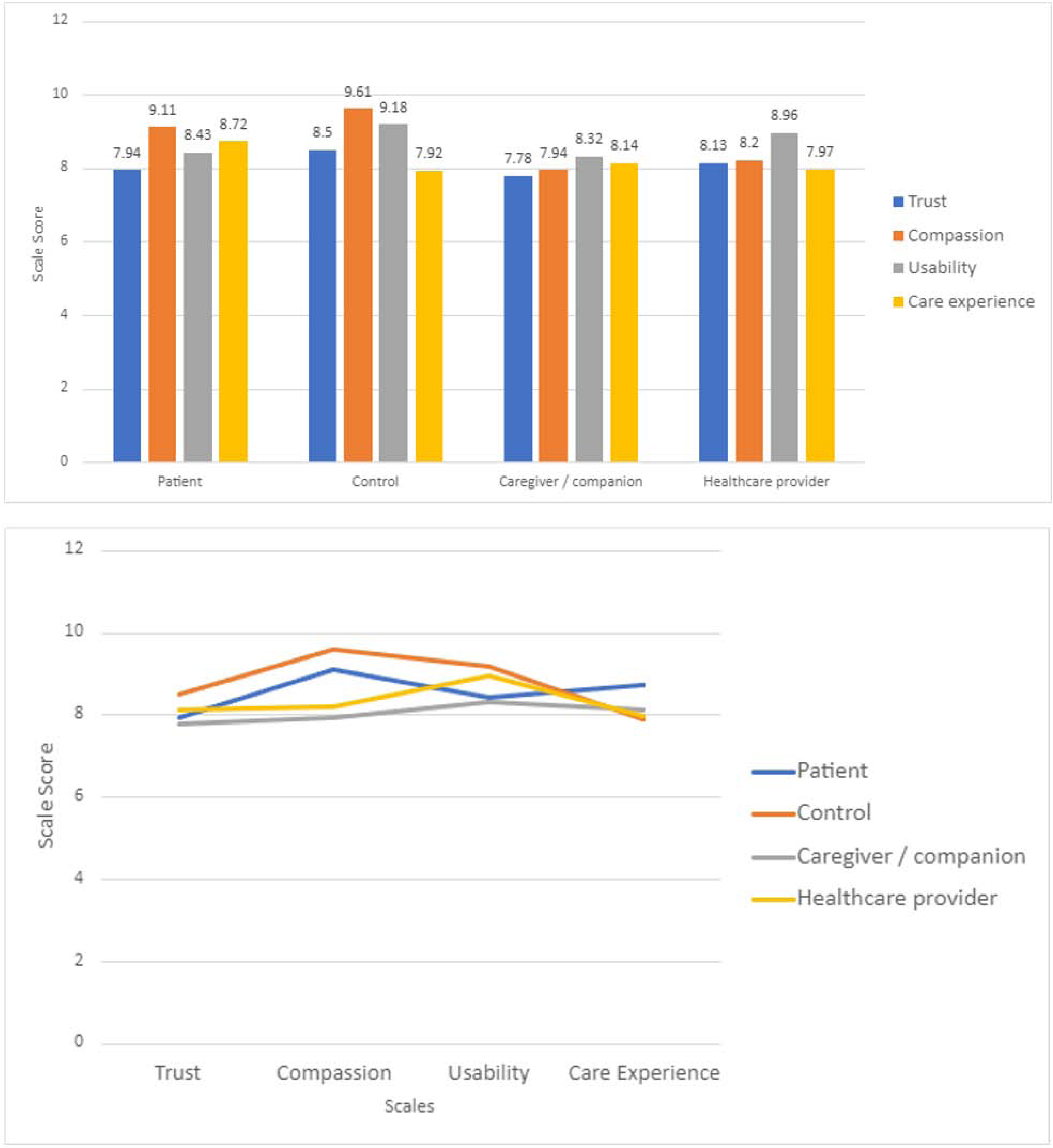
Mean scores on scales within each participant category.

### Effects of scale by participant category

The results of the repeated measure ANOVA indicated that there was no overall statistically significant interaction between scale and participant category, indicating that the difference between the 4 scales is consistent within each participant category, with large effect size (Table 3).

### Effects of sex

The results of the repeated measure ANOVA indicated that there was no overall statistically significant difference of mean survey scores between the Male and Female gender categories, with medium effect size (Table 4). That is, Males and Females scored similarly on the overall survey (Figure 5).

**Figure 5.**
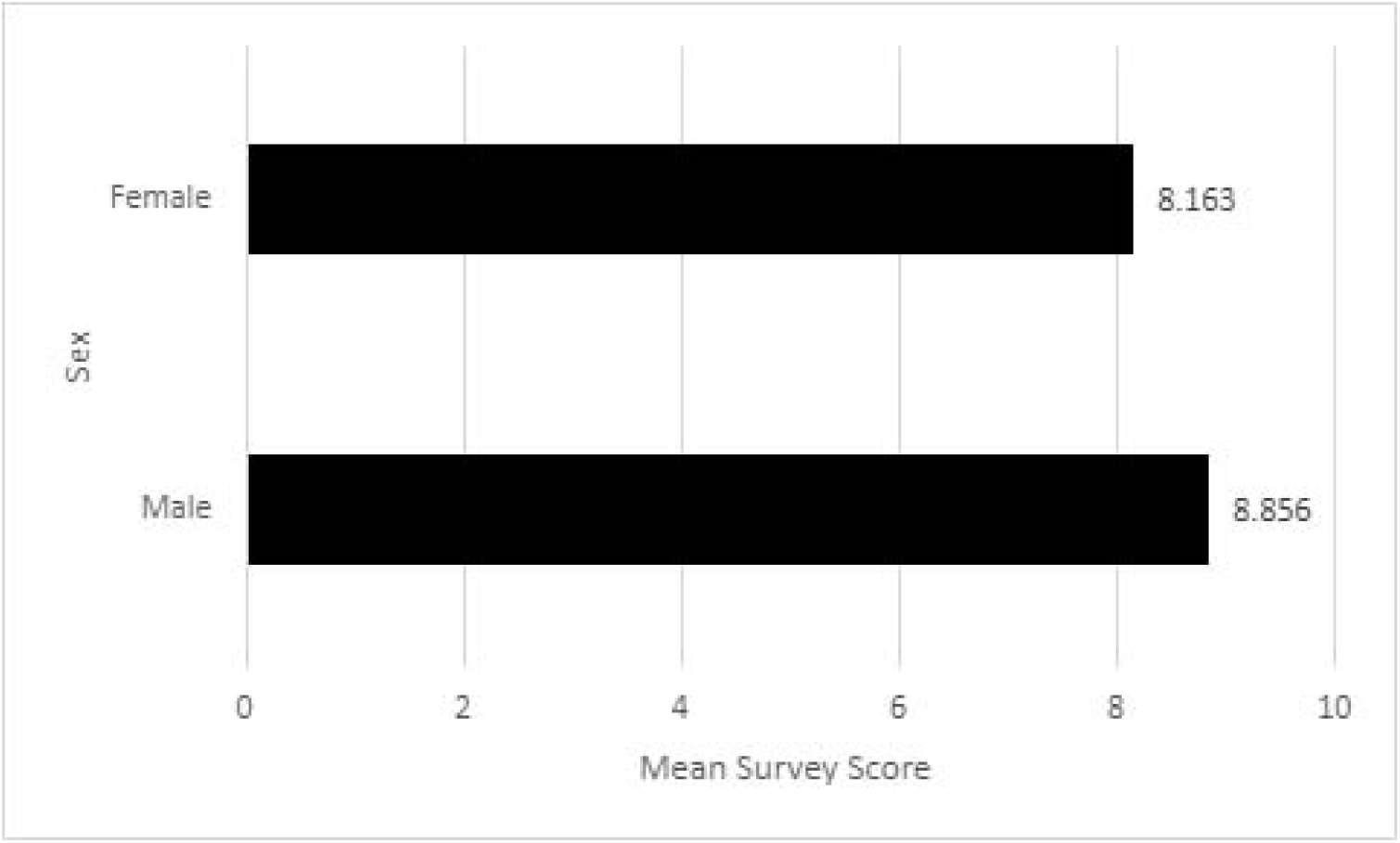
Overall mean survey scores of Males and Females.

**Table 4.** Results of the ANOVA of Scale by Sex.

| | F | df | Sig. (p) | $\eta^2$ |
| --- | --- | --- | --- | --- |
| Sex | 2.239 | 1,21 | 0.149 | 0.096 |
| Scale * Sex | 0.457 | 3,63 | 0.713 | 0.021 |

Note: Scale (Trust, Compassion, Usability, Care Experience)
<sup>z</sup>Eta squared: small es .01-.058; medium es .059-.137; large es >.137**Figure 5***Overall mean survey scores of Males and Females***Figure 6***Mean scores on scales of Males and Females****Effects of device type***

The results of the repeated measure ANOVA indicated that there was no overall statistically significant interaction between scale and sex, indicating that the difference between the 4 scales is consistent within each sex category, with large effect size (Table 4). That is, the sex did not significantly affect a participant’s performance on any of the 4 scales (Figure 6).

**Figure 6.**
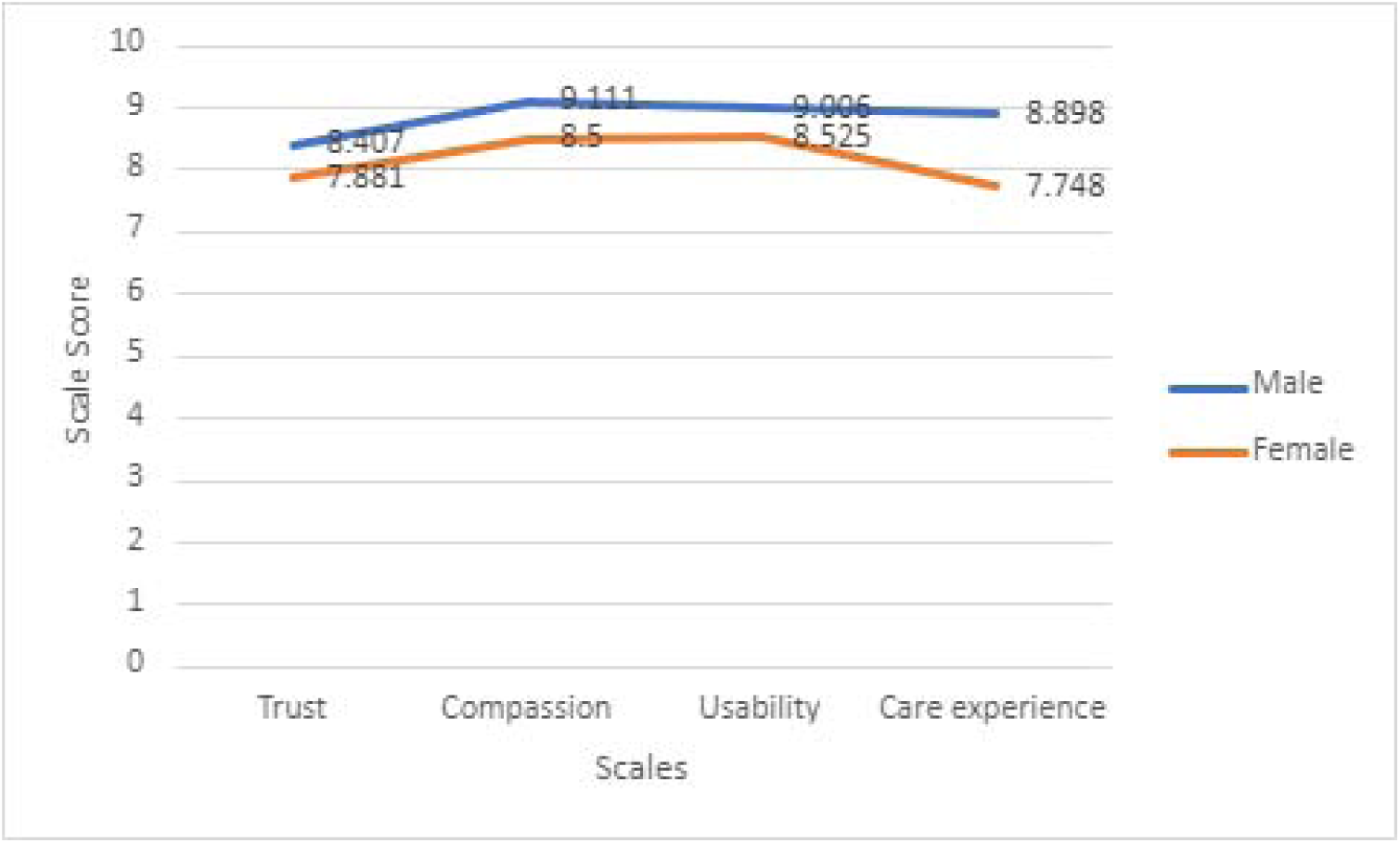
Mean scores on scales of Males and Females.

### Effects of device type

The results of the repeated measure ANOVA indicated that there was no overall statistically significant difference of mean survey scores between the types of devices used, with large effect size (Table 5). That is, the type of device used (Figure 7) did not significantly affect the overall survey scores (Figure 8).

**Figure 7.**
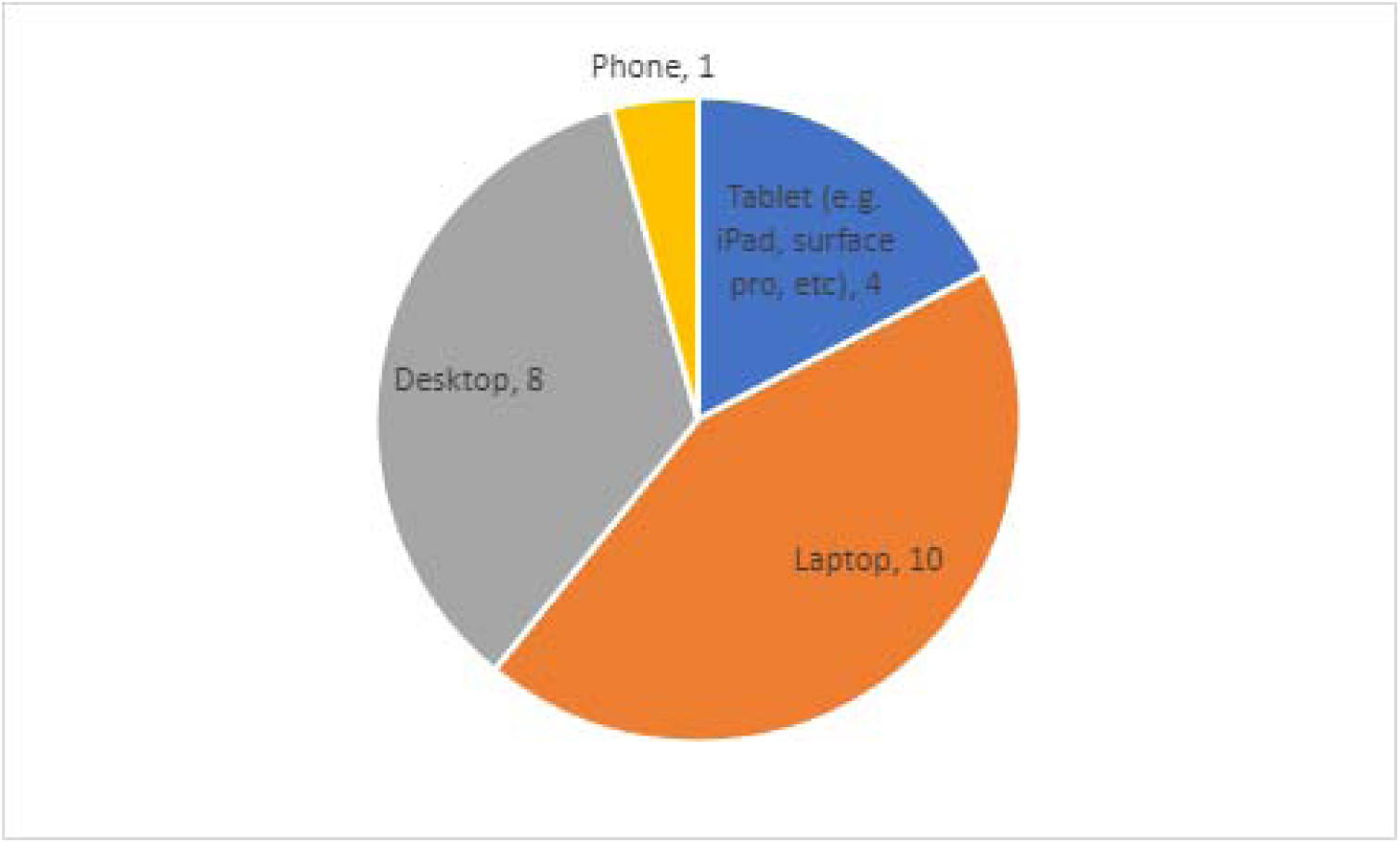
Types of devices used by participants during the assessment.

**Figure 8.**
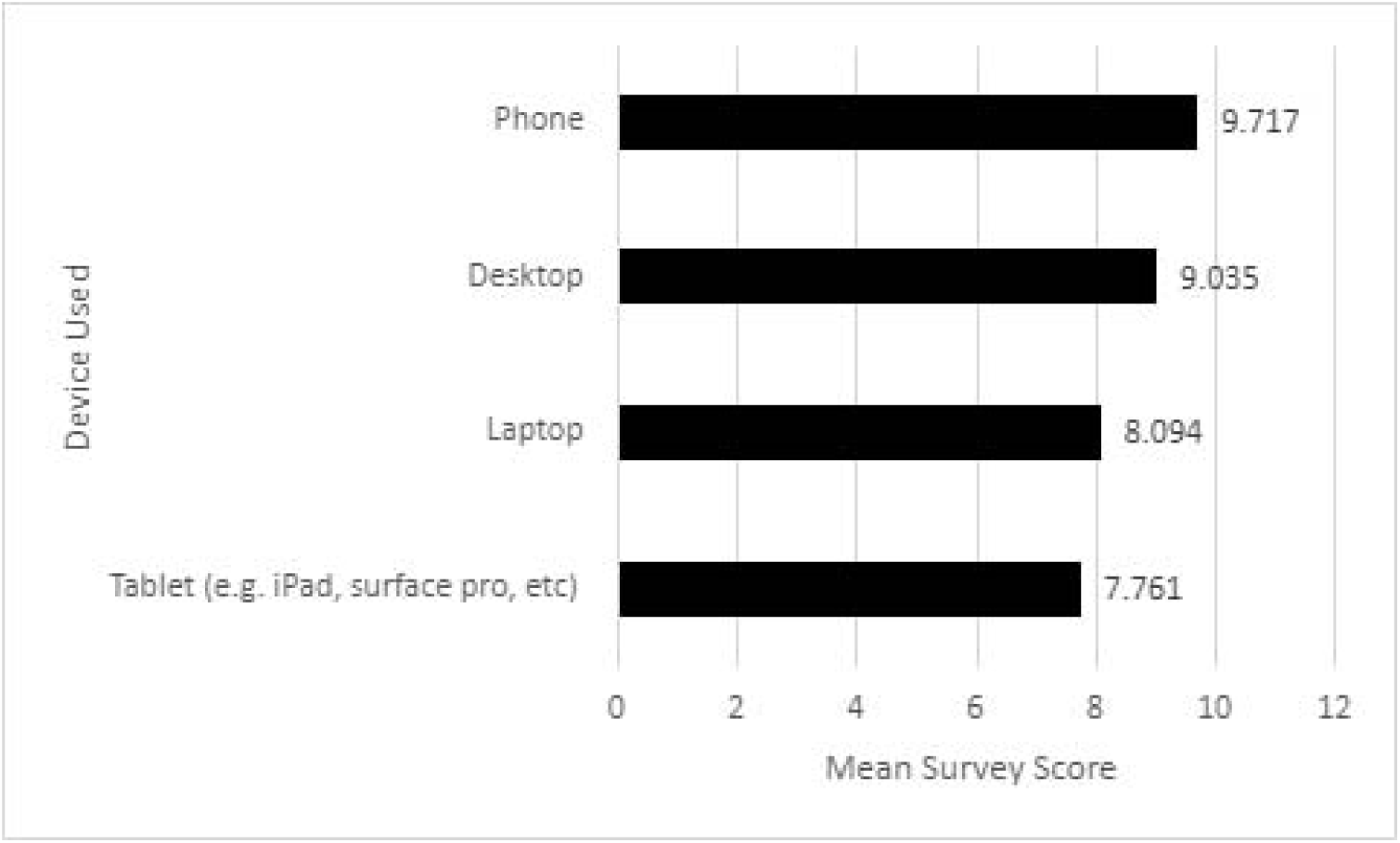
Mean Survey Score by Device Type.

**Table 5.** Results of the ANOVA of Scale by Device Type.

| | F | df | Sig. (p) | $\eta^2$ |
| --- | --- | --- | --- | --- |
| Device Type | 2.407 | 3,19 | 0.099 | 0.275 |
| Scale * Device Type | 0.247 | 9,57 | 0.985 | 0.038 |

Note: Scale (Trust, Compassion, Usability, Care Experience)
<sup>z</sup>Eta squared: small es .01-.058; medium es .059-.137; large es >.137

The results of the repeated measure ANOVA indicated that there was no overall statistically significant interaction between scale and device type, indicating that the difference between the 4 scales is consistent within each device type, with small effect size (Table 5). That is, the device type did not significantly affect a participant’s performance on any of the 4 scales (Figure 9).

**Figure 9.**
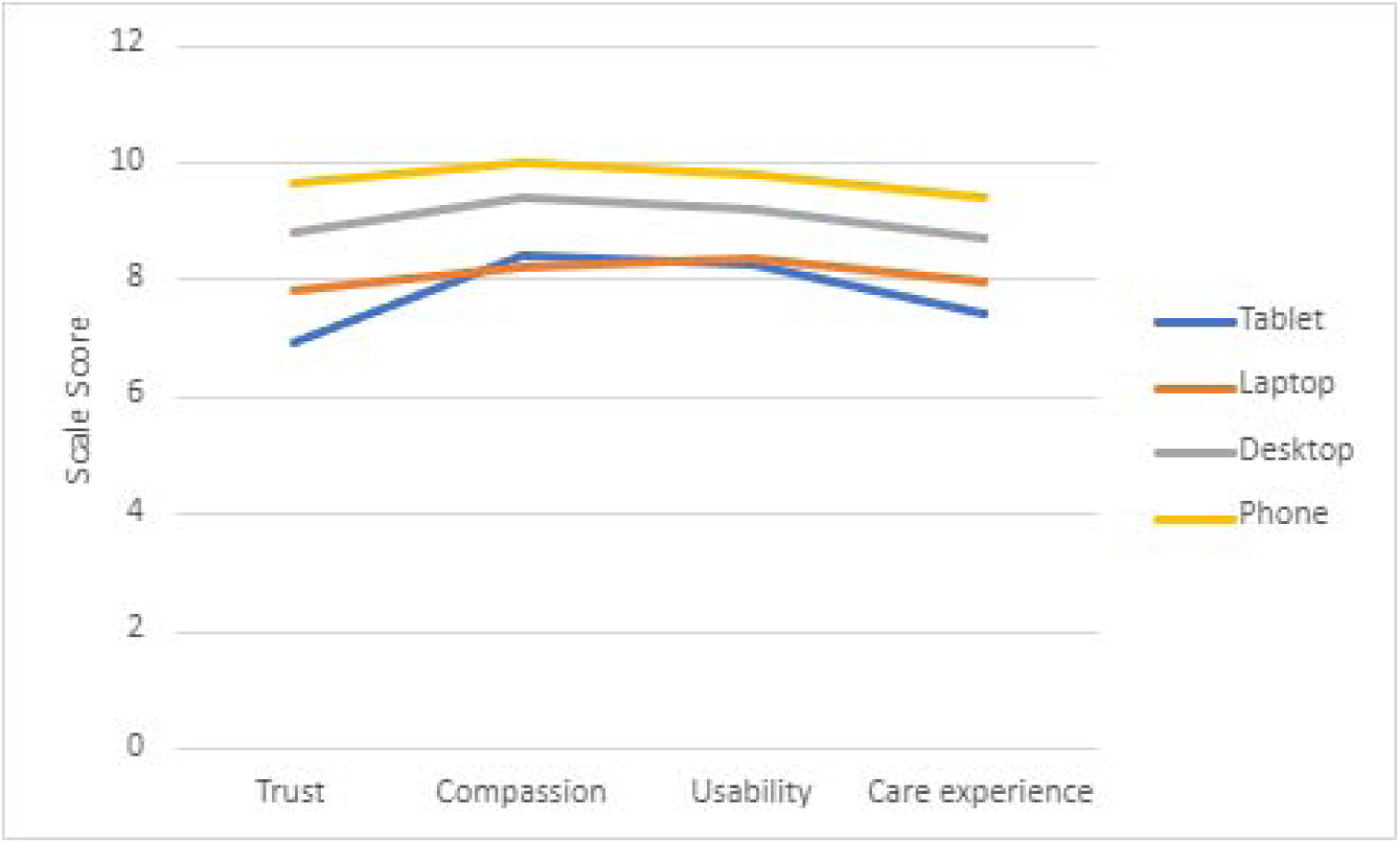
Mean scores on Scales by Device Type.

### Effects of chatbot use

The results of the repeated measure ANOVA indicated that there was no overall statistically significant difference of mean survey scores between those who used and did not use the chatbot, with small effect size (Table 6). That is, those who used and didn’t use the chatbot scored similarly on the survey (Figure 10).

**Figure 10.**
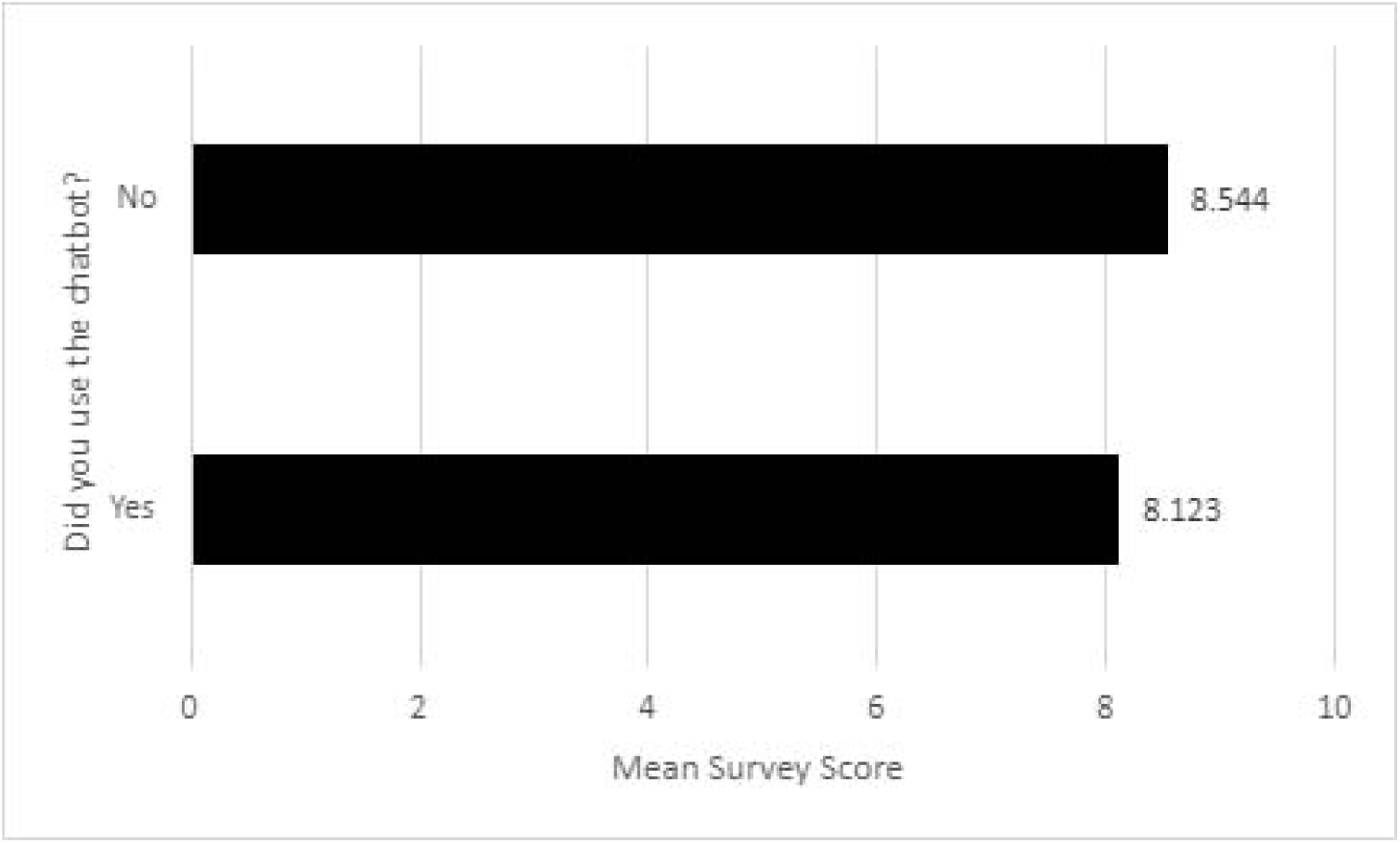
Mean survey scores by chatbot usage.

**Table 6.**
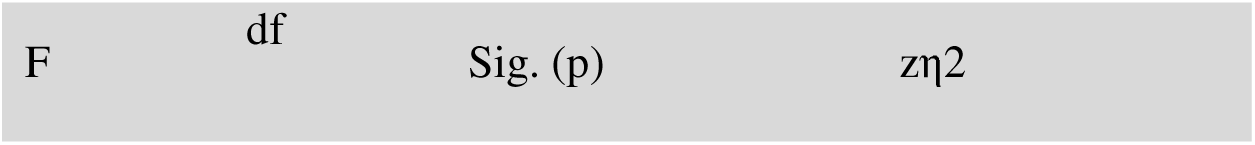
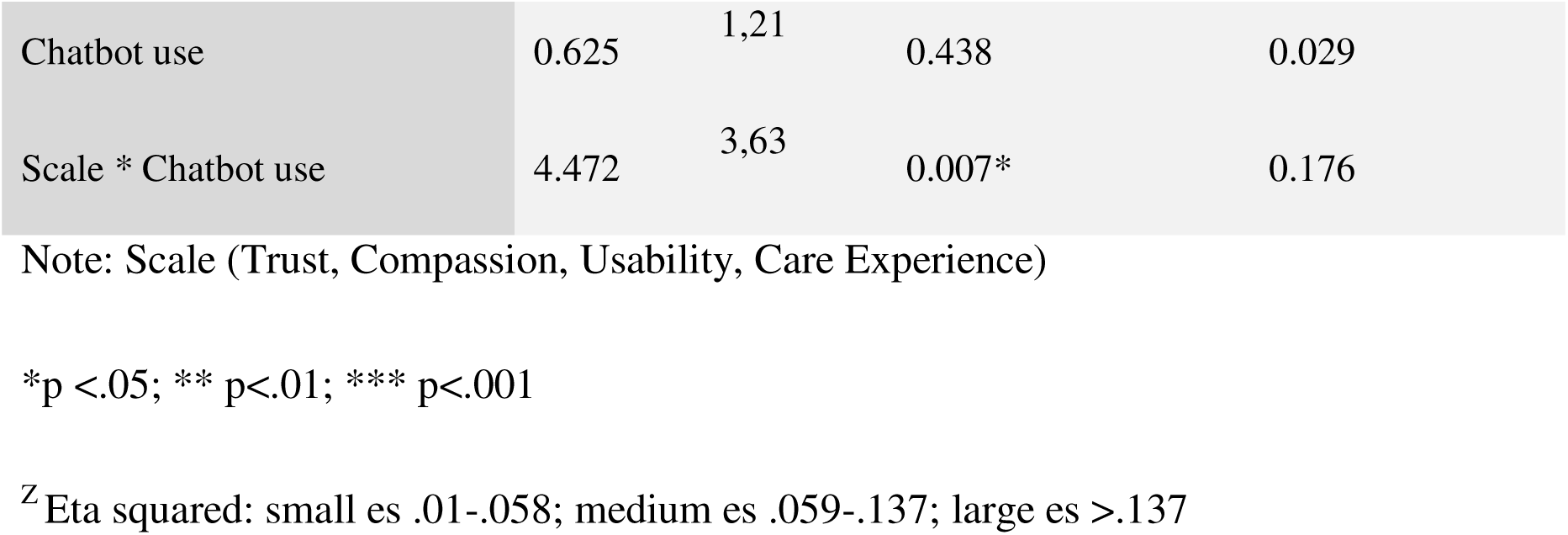
Results of the ANOVA of Scale by Chatbot use.

| F | df | Sig. (p) | $\eta^2$ |
| --- | --- | --- | --- |

|  |  |  |  |  |
| --- | --- | --- | --- | --- |
| Chatbot use | 0.625 | 1,21 | 0.438 | 0.029 |
| Scale * Chatbot use | 4.472 | 3,63 | 0.007* | 0.176 |

Note: Scale (Trust, Compassion, Usability, Care Experience)
\*p <.05; \*\* p<.01; \*\*\* p<.001
<sup>z</sup>Eta squared: small es .01-.058; medium es .059-.137; large es >.137

The results of the repeated measure ANOVA indicated that there was a significant interaction between scale and chatbot use, indicating that the difference between the 4 scales is significant when considering chatbot use, with large effect size (Table 6). That is, using the chatbot led to significant differences in the scores on the 4 scales (Figure 11). More specifically, those who used the chatbot feature during the assessment scored close to 2 whole points lower on the Usability scale compared to those who did not use the chatbot feature (Figure 11). It should be noted that one participant attempted to use the chatbot feature but was unable due to confusion over how to operate it.

**Figure 11.**
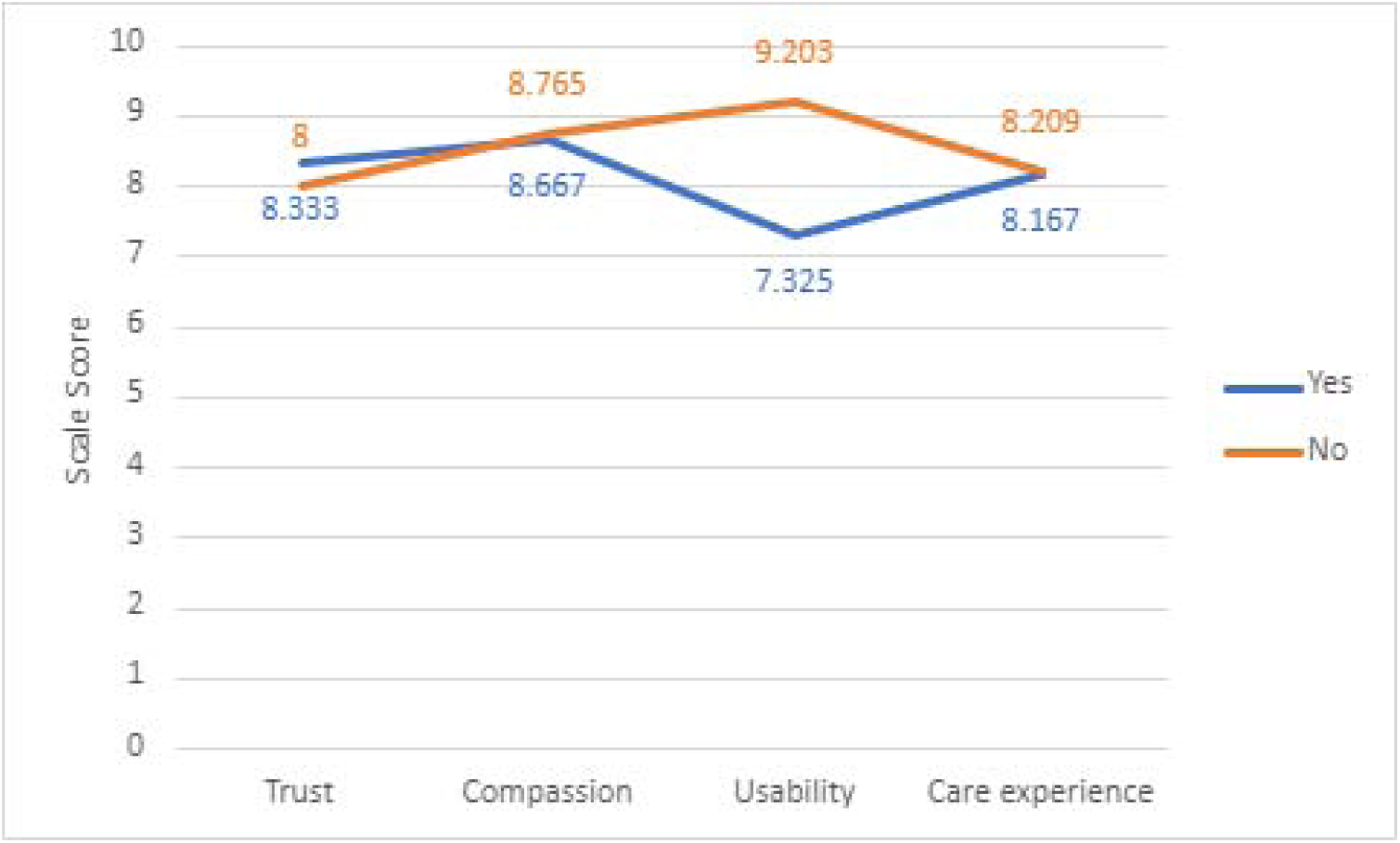
Mean scores on Scales by chatbot use.

### Effects of chatbot familiarity

The results of the repeated measure ANOVA indicated that there was no overall statistically significant difference of mean survey scores between the different levels of chatbot familiarity, with large effect size (Table 7). That is, participants with varying levels of chatbot familiarity (Figure 12) scored similarly on the survey (Figure 13).

**Figure 12.**
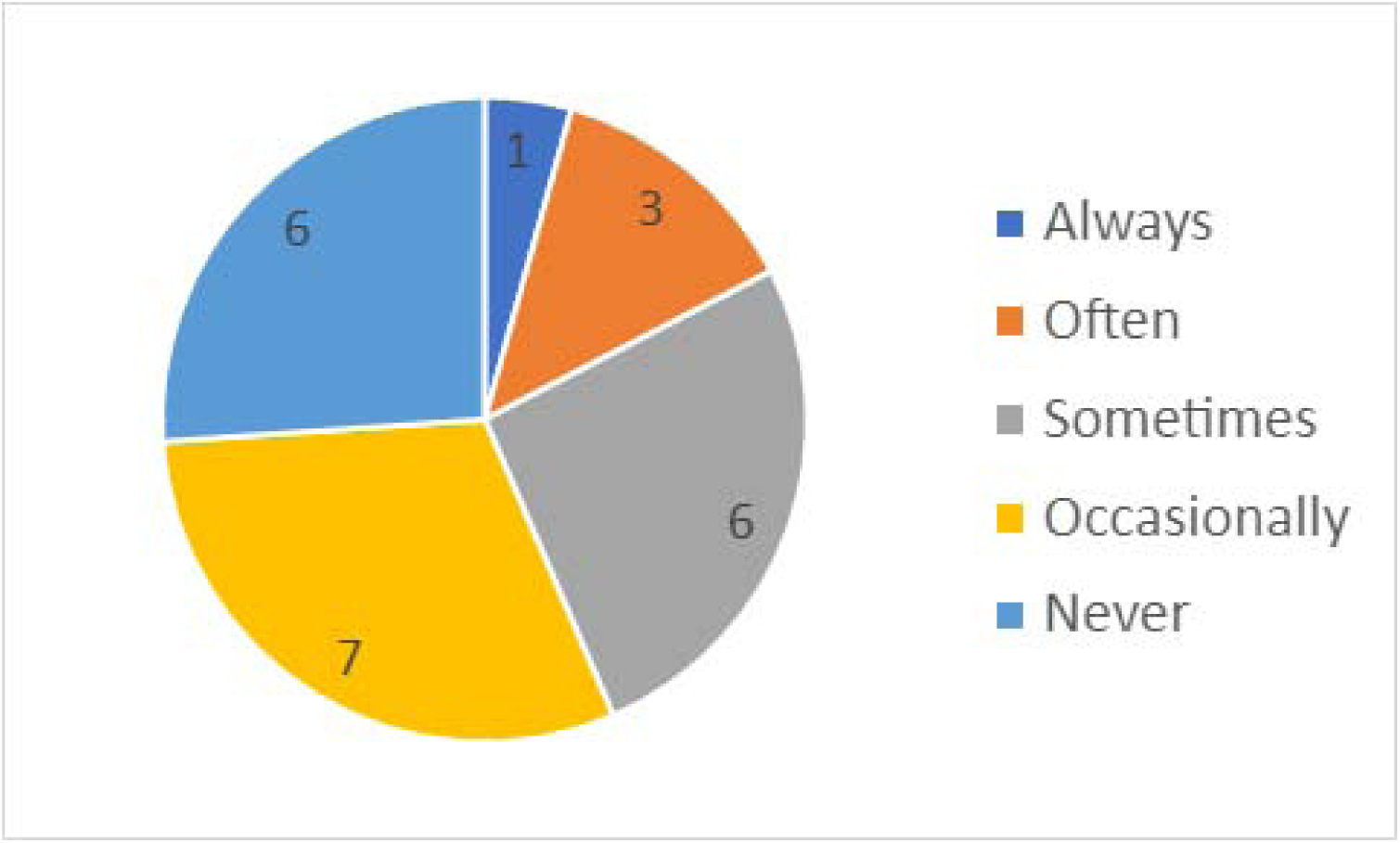
Chatbot technology (Google assistant, Alexa, Siri etc.) usage levels among participants.

**Figure 13.**
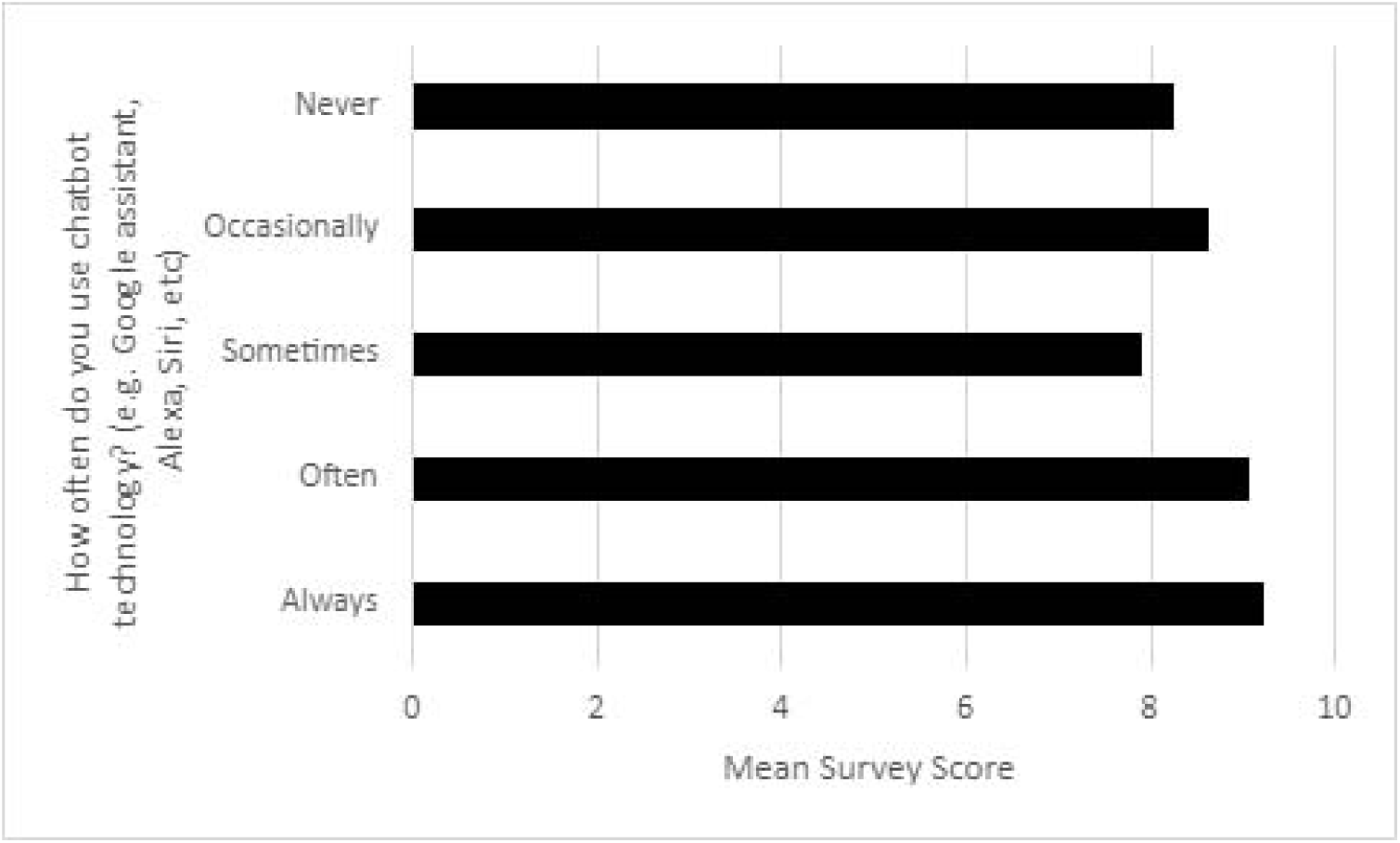
Mean survey scores by chatbot technology familiarity.

**Table 7.** Results of the ANOVA of Scale by Chatbot familiarity.

| | F | df | Sig. (p) | $\eta^2$ |
| --- | --- | --- | --- | --- |
| Chatbot familiarity | 0.759 | 4,18 | 0.565 | 0.144 |
| Scale * Chatbot familiarity | 0.556 | 12,54 | 0.867 | 0.11 |

Note: Scale (Trust, Compassion, Usability, Care Experience)
<sup>z</sup>Eta squared: small es .01-.058; medium es .059-.137; large es >.137**Figure 12***Chatbot technology (Google assistant, Alexa, Siri etc.) usage levels among participants***Figure 13***Mean survey scores by chatbot technology familiarity***Figure 14***Mean scores on Scales by chatbot familiarity****Effects of Education Level***

The results of the repeated measure ANOVA indicated that there was no statistically significant interaction between scale and chatbot familiarity, indicating that the difference between the 4 scales is consistent within the different levels of chatbot familiarity, with medium effect size (Table 7). That is, participants with varying levels of chatbot familiarity scored similarly in each of the 4 scales (Figure 14).

**Figure 14.**
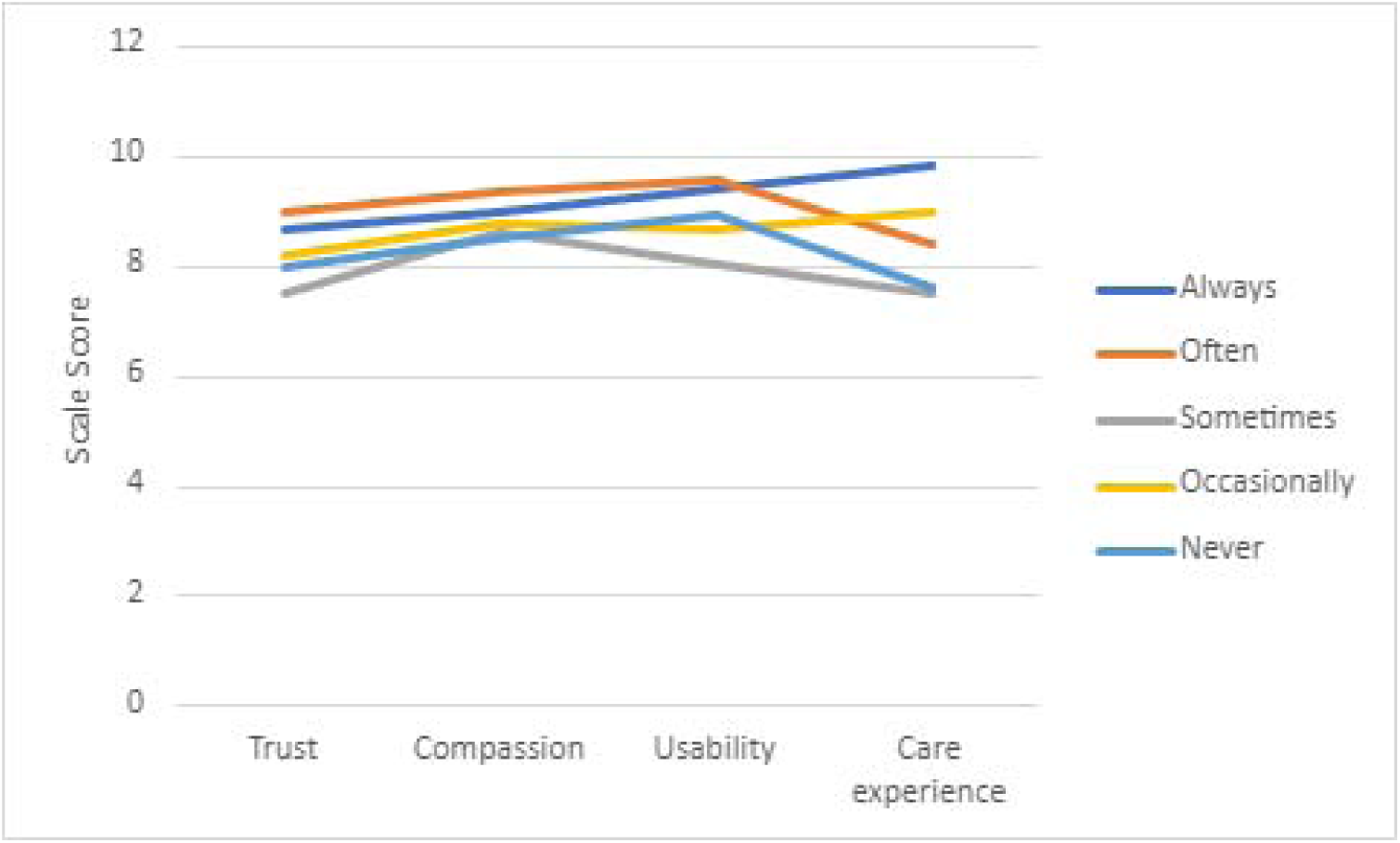
Mean scores on Scales by chatbot familiarity.

### Effects of Education Level

The results of the repeated measure ANOVA indicated that there was no overall statistically significant difference of mean survey scores between the different levels of education, with large effect size (Table 8). That is, a participant’s education level (Figure 15) did not significantly affect the overall survey scores (Figure 16).

**Figure 15.**
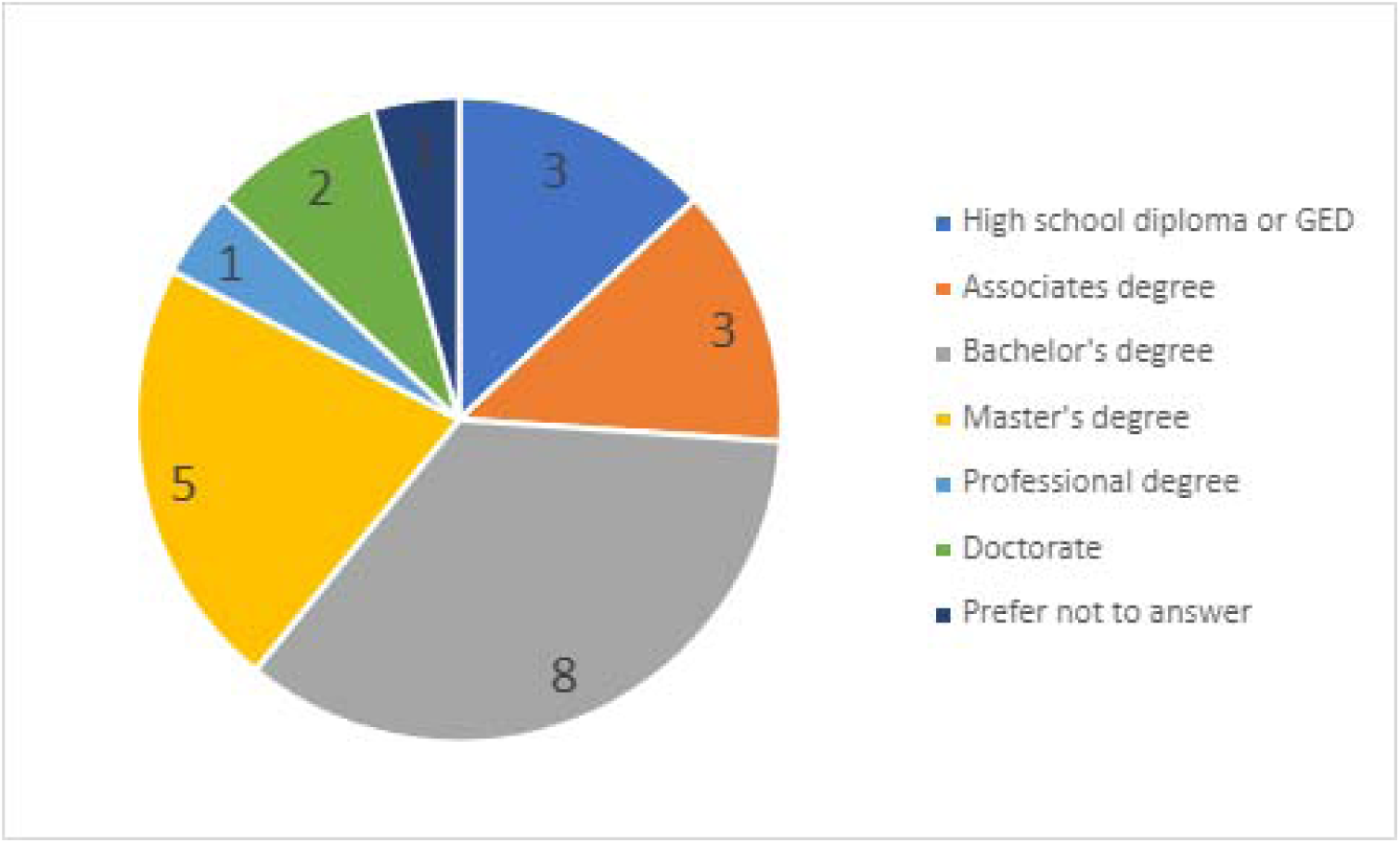
Education levels among participants.

**Figure 16.**
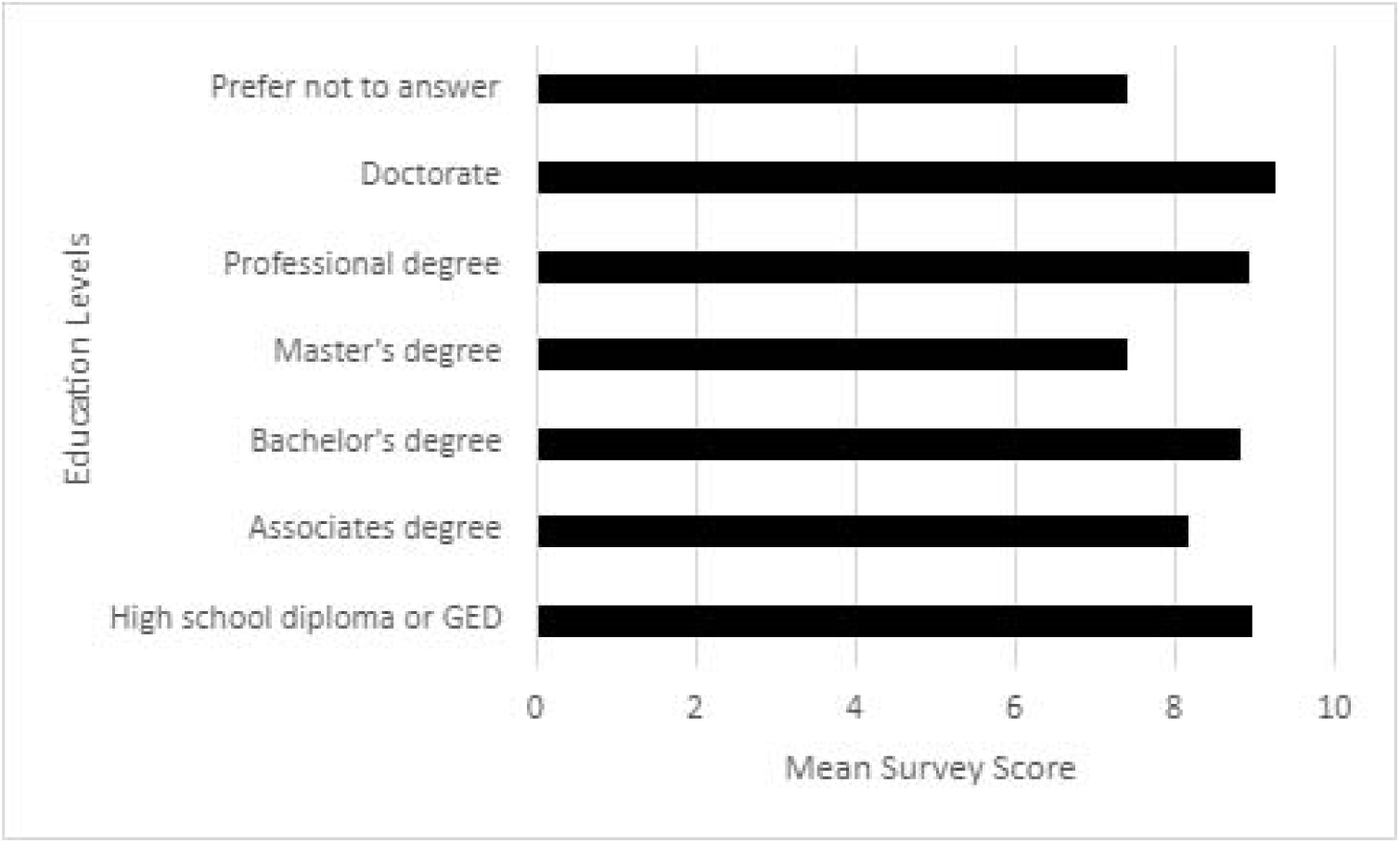
Mean survey scores by education level.

**Table 8.** Results of the ANOVA of Scale by Education Level.

| | F | df | Sig. (p) | $\eta^2$ |
| --- | --- | --- | --- | --- |
| Education Level | 1.603 | 6,16 | 0.21 | 0.375 |
| Scale * Education Level | 1.776 | 18,48 | 0.058 | 0.4 |

Note: Scale (Trust, Compassion, Usability, Care Experience)
<sup>z</sup>Eta squared: small es .01-.058; medium es .059-.137; large es >.137

The results of the repeated measure ANOVA indicated that there was no statistically significant interaction between scale and education level, indicating that the difference between the 4 scales is consistent within the different levels of education level, with large effect size (Table 8). That is, participants with varying levels of education level scored similarly on each of the 4 scales (Figure 17).

**Figure 17.**
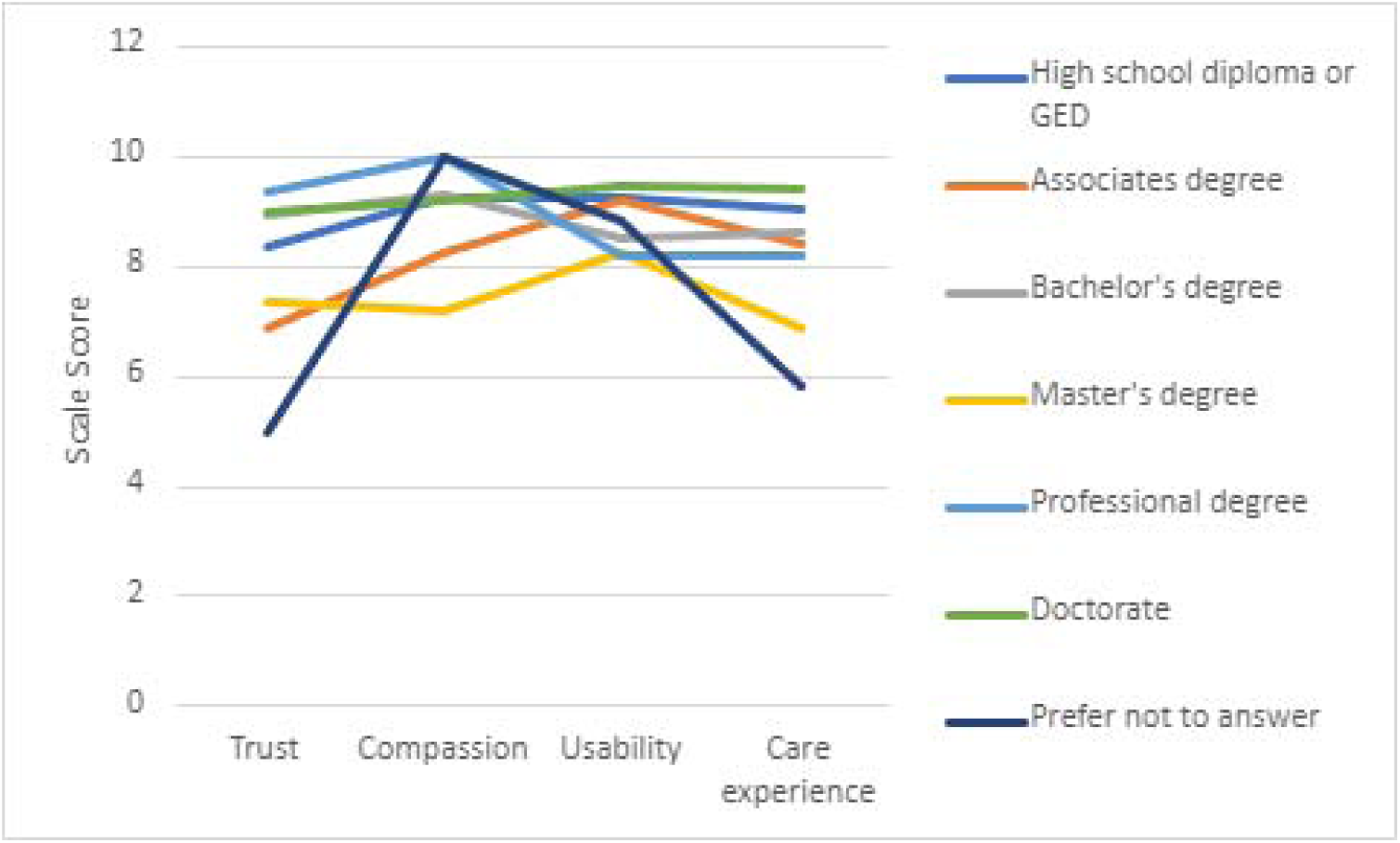
Mean scores on Scales by Education Level.

### Effects of cognitive assessment administration

The results of the repeated measure ANOVA indicated that there was no overall statistically significant difference of mean survey scores among healthcare providers when considering the number of cognitive assessments administered, with medium effect size (Table 9). That is, the number of cognitive assessments that a healthcare provider has administered in their careers (Figure 18) did not significantly affect the overall survey scores (Figure 19).

**Figure 18.**
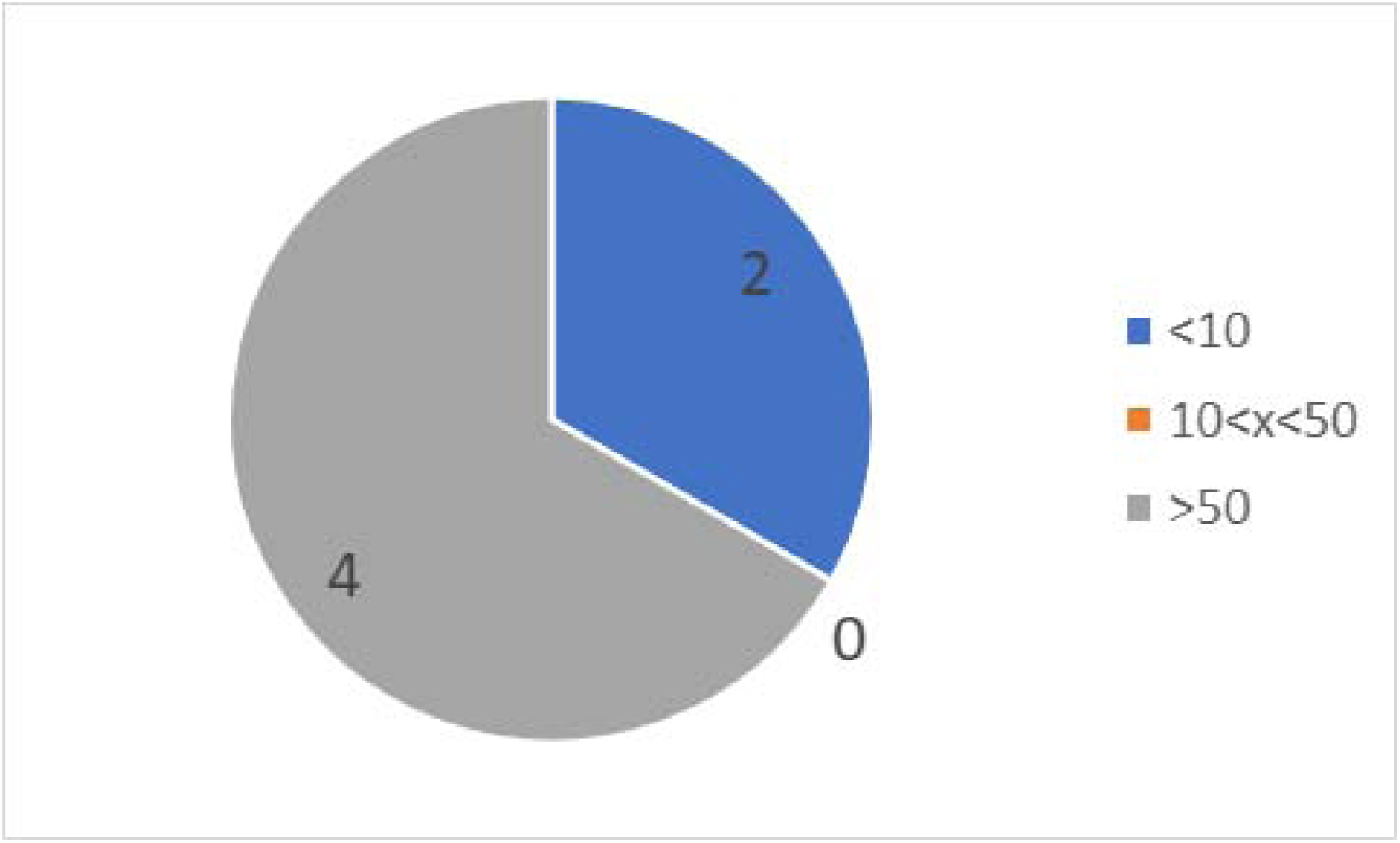
Number of cognitive assessments administered among healthcare providers.

**Figure 19.**
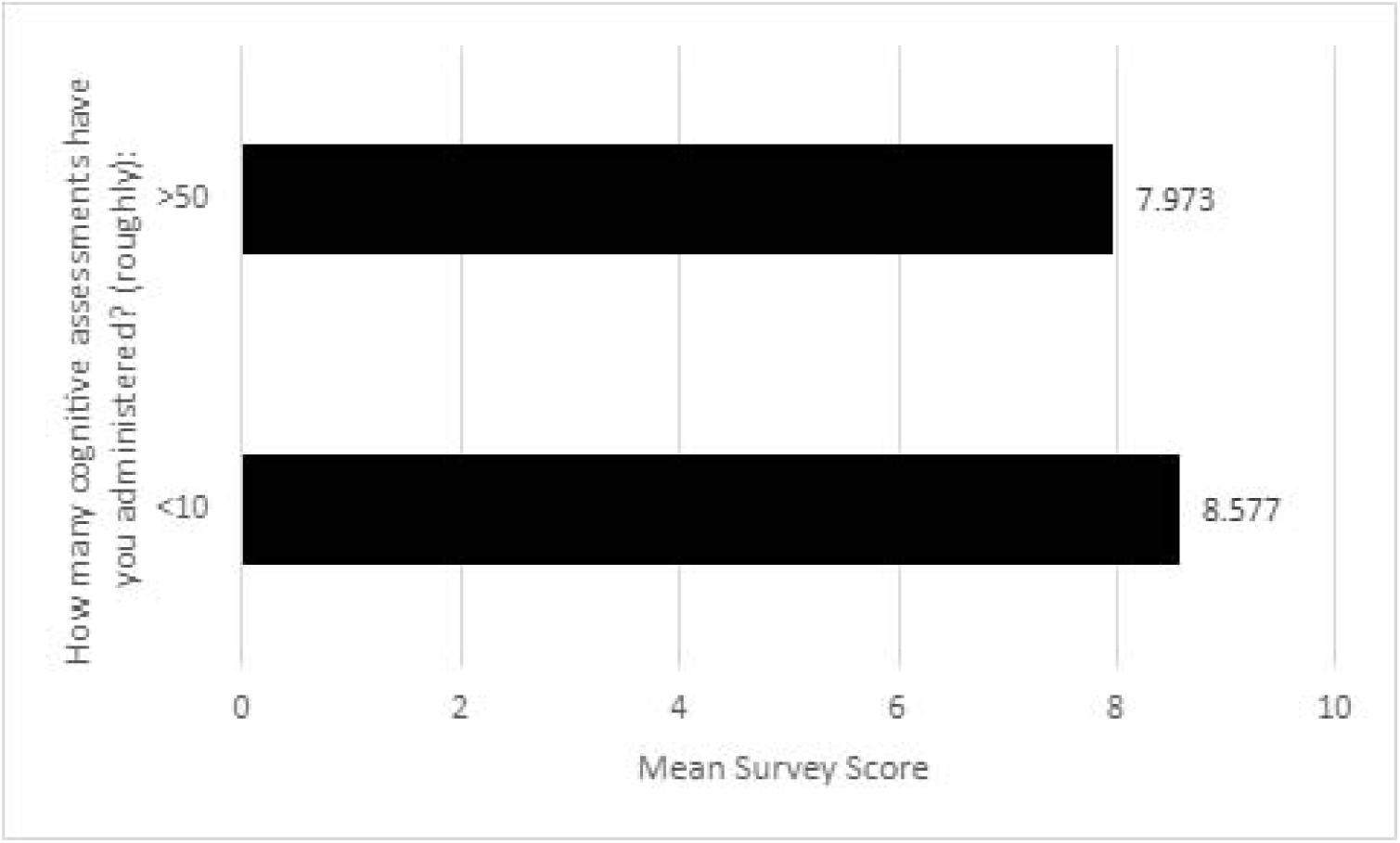
Mean survey scores by number of cognitive assessments administered among healthcare providers.

**Table 9.** Results of the ANOVA of Scale by Cognitive Assessment Administration.

| | F | df | Sig. (p) | $\eta^2$ |
| --- | --- | --- | --- | --- |
| Cognitive Assessment Administration | 0.329 | 4,18 | 0.597 | 0.076 |
| Scale * Cognitive Assessment Administration | 0.09 | 12,54 | 0.964 | 0.022 |

Note: Scale (Trust, Compassion, Usability, Care Experience)
<sup>Z</sup> Eta squared: small es .01-.058; medium es .059-.137; large es >.137

The results of the repeated measure ANOVA indicated that there was no statistically significant interaction between scale and cognitive assessment administration indicating that the difference between the 4 scales is consistent within the different levels of education level, with small effect size (Table 9). That is, the number of cognitive assessments administered by a healthcare provider did not amount to significant differences in the scores on each of the 4 scales (Figure 20).

**Figure 20.**
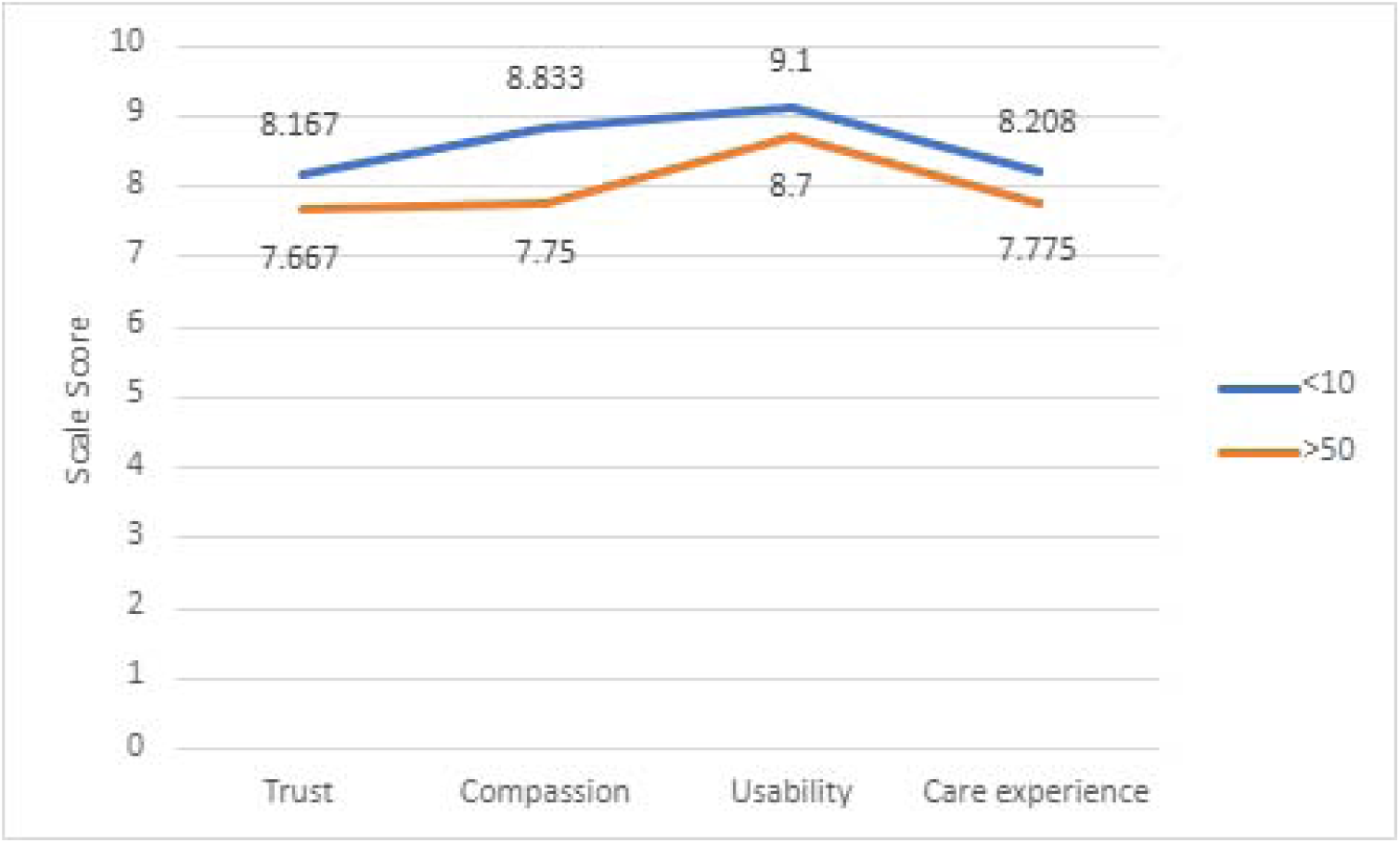
Mean scores on Scales by number of cognitive assessments administered among healthcare providers.

### Effects of caregiving experience

The results of the repeated measure ANOVA indicated that there was no overall statistically significant difference of mean survey scores among caregivers when considering the number of years of caregiving experience with large effect size (Table 10). That is, the number of years of caregiving experience (Figure 21) did not significantly affect the overall survey scores (Figure 22).

**Figure 21.**
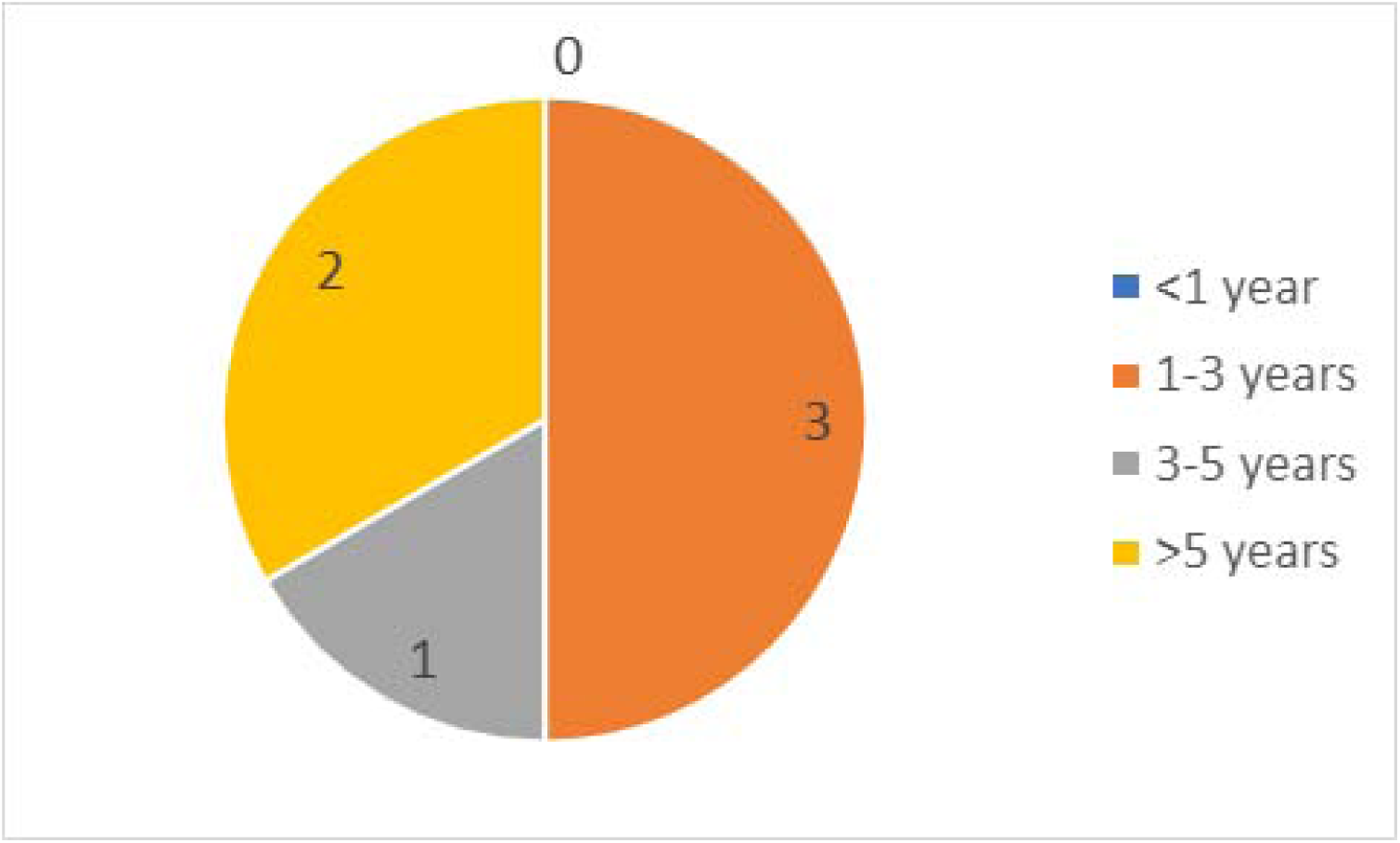
Number of caregiver years among caregivers.

**Figure 22.**
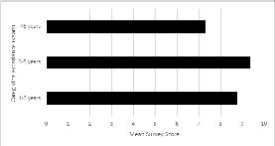
Mean survey scores by caregiving experience among caregivers.

**Table 10.** Results of the ANOVA of Scale by Caregiving years.

| | F | df | Sig. (p) | $\eta^2$ |
| --- | --- | --- | --- | --- |
| Number of caregiving years | 1.01 | 2,4 | 0.441 | 0.336 |
| Scale * Number of caregiving years | 0.974 | 6,12 | 0.483 | 0.327 |

Note: Scale (Trust, Compassion, Usability, Care Experience)
<sup>z</sup> Eta squared: small es .01-.058; medium es .059-.137; large es >.137

The results of the repeated measure ANOVA indicated that there was no statistically significant interaction between scale and caregiving experience indicating that the difference between the 4 scales is consistent within the different levels of caregiving experience, with large effect size (Table 10). That is, the number of years of caregiving experience did not amount to significant differences in the scores on each of the 4 scales (Figure 23).

**Figure 23.**
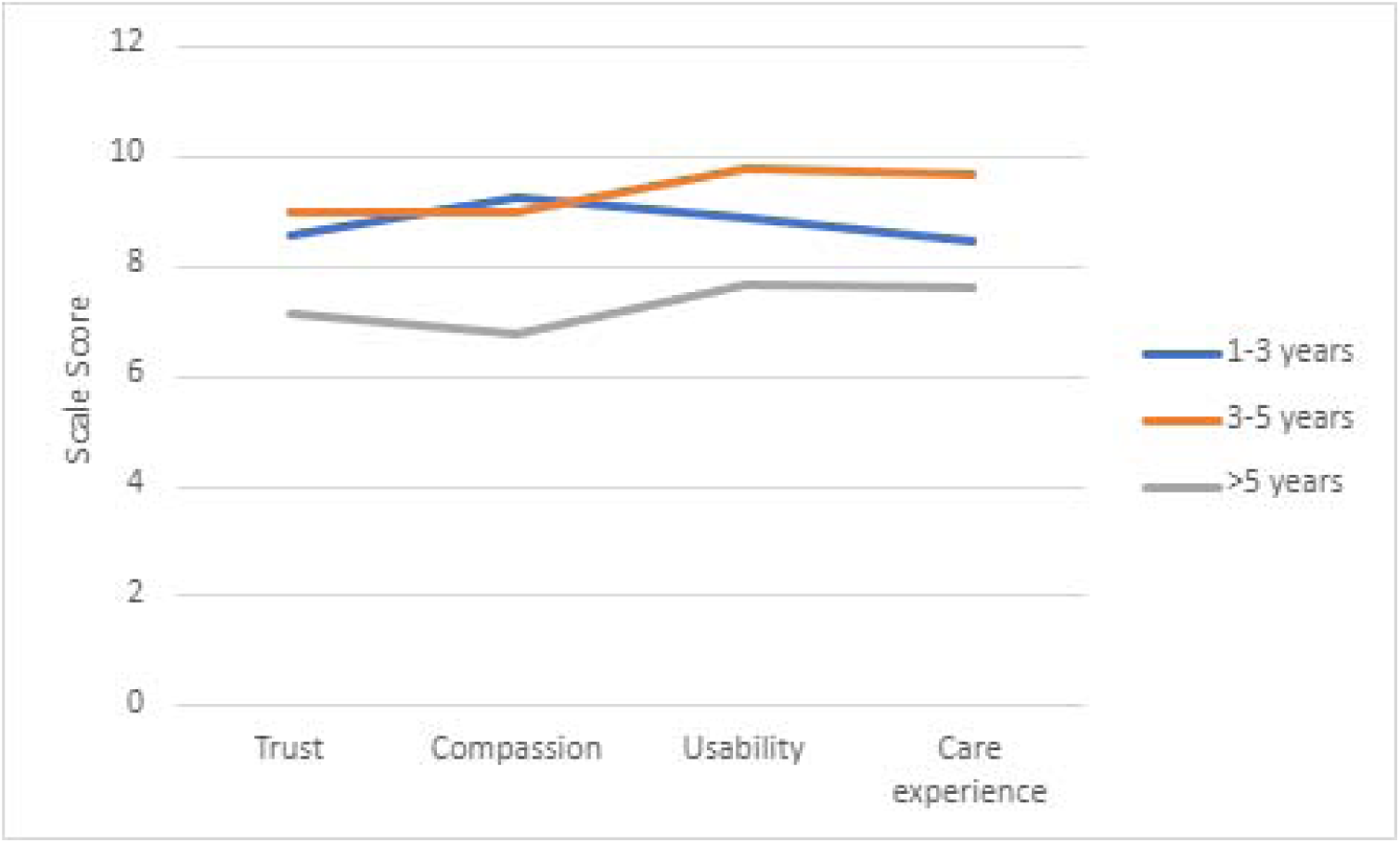
Mean scores on Scales by Caregiving experience.

### Effect Size

Although many of our results did not find statistically significant findings, the effect size did indicate that differences did indeed exist in many of the analyzed variables (Table 11). For between-subject effects, a medium or large effect size indicates that differences exist between the variable of interest and the mean survey score. For example, when looking at Device Type, a large effect size indicates that differences exist in survey scores based on what kind of device participants chose to use for the screen. For within-subject effects, a medium or large effect size indicates that differences exist between the 4 scales within each sub-group of the variable of interest. For example, when looking at Scale*Participant Category, a large effect size indicates that differences exist between the 4 scales in each of the patient, caregiver, healthcare provider, and control participant groups.

**Table 11.**
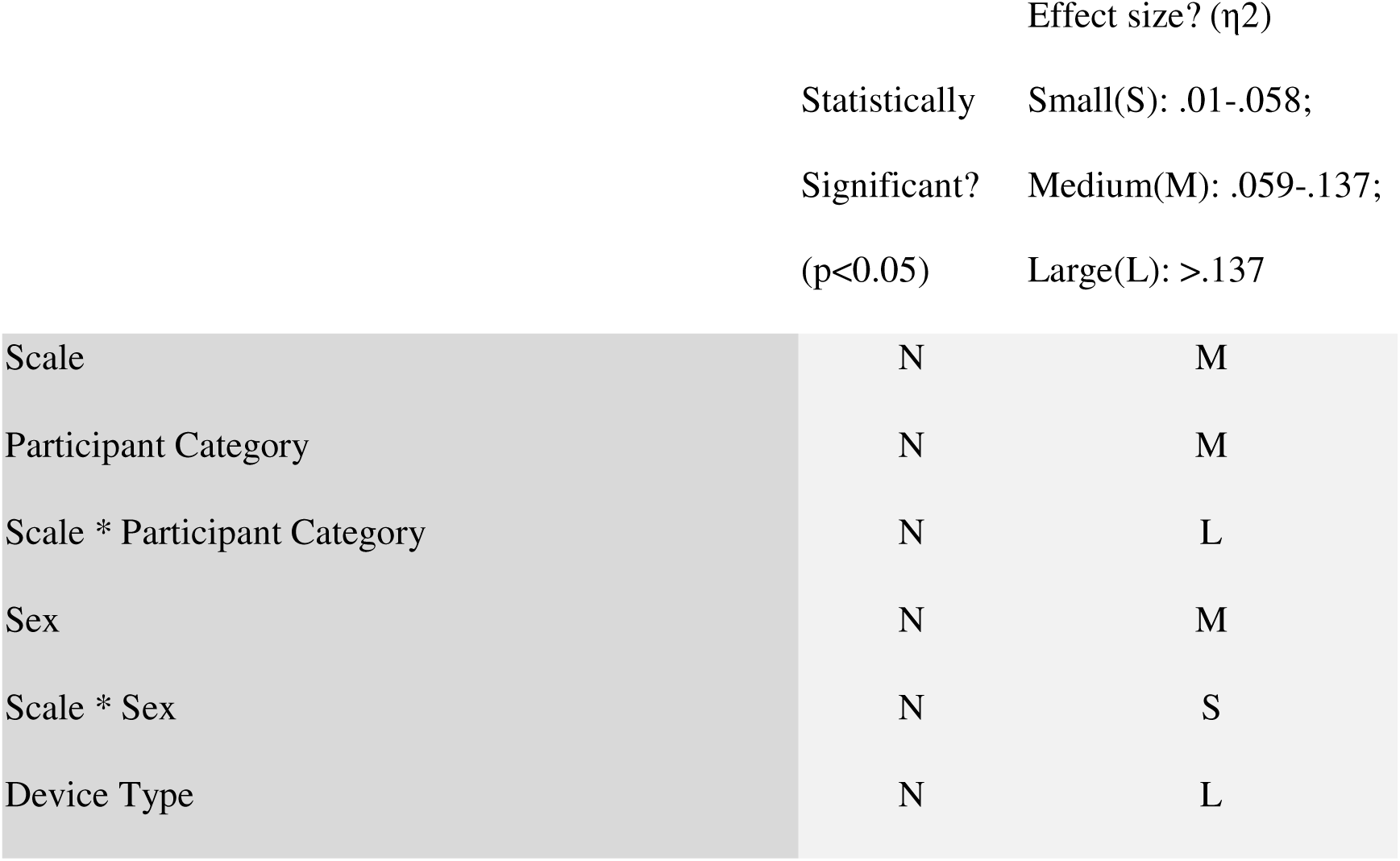
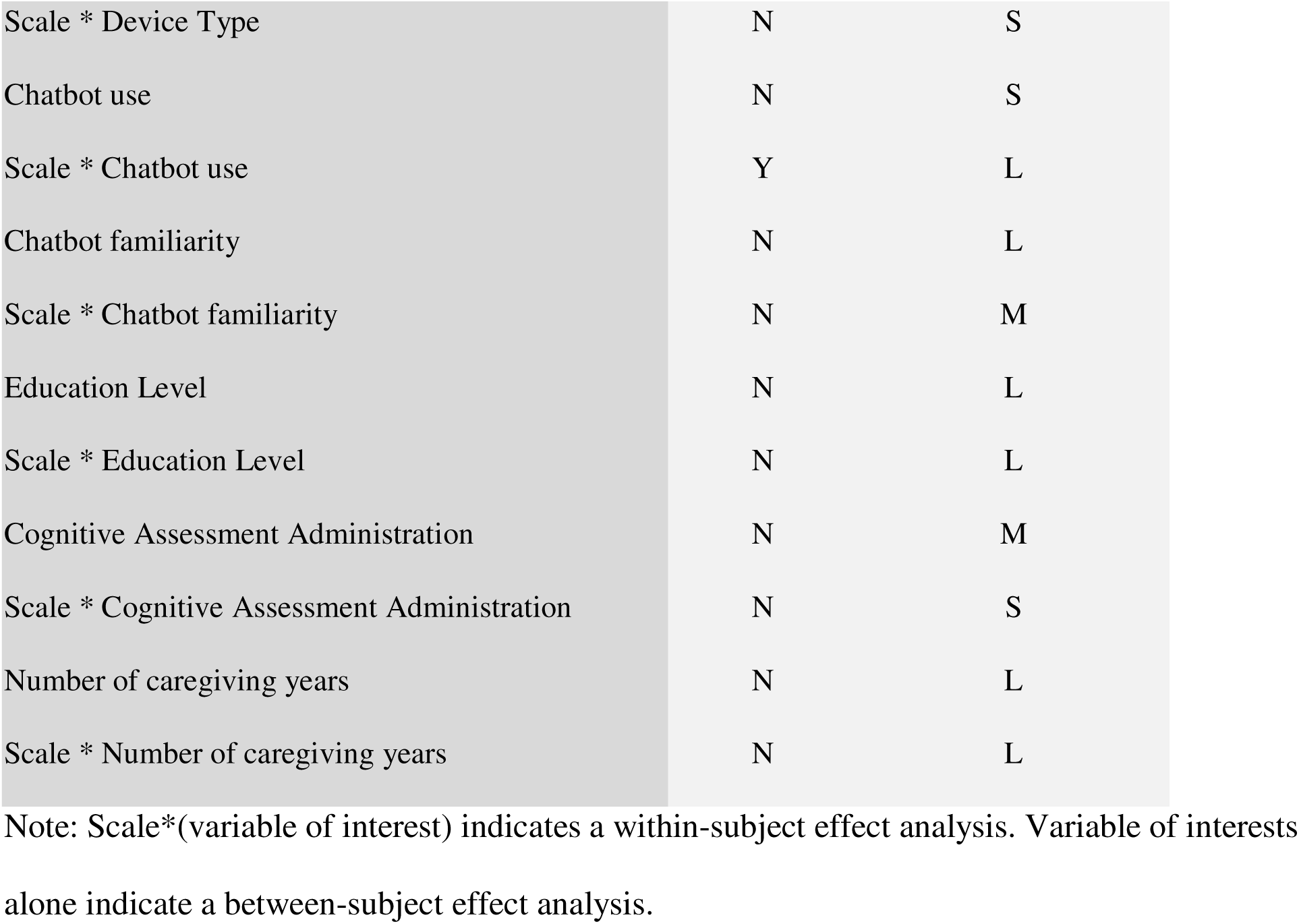
Summary of statistical significance and effect size.

## Discussion

The findings indicate that there were no statistically significant differences between the overall mean survey values of each participant category. In other words, patients, controls, caregivers, and healthcare providers all scored similarly on the overall survey. Additionally, within each participant group, participants scored similarly across all 4 scales: Trust, Compassion, Usability, and Care Experience. This suggests that the assessment was experienced similarly by each group and that the assessment did not bias a particular group such as cognitively impaired users. There was no statistically significant difference between the overall survey scores of healthcare providers when considering the number of cognitive assessments they have administered. Additionally, there was no statistically significant difference between the overall survey scores of caregivers when considering the number of years of caregiving experience. In other words, healthcare providers and caregivers experienced the assessment similarly regardless of the number of cognitive assessments or the number of caregiving years provided. Factors such as sex, device type, chatbot familiarity, and level of education had no statistically significant effect on overall survey or scale-specific scores, suggesting that the assessment did not significantly bias a specific group based on demographic attributes.

One statistically significant difference observed was between the mean scores of the scales Trust and Compassion. Participants, regardless of what category they were in, scored significantly higher on questions related to Compassion compared to questions related to Trust on the survey. The overall mean score for the scale Trust was 8.09 while the overall mean score for the scale Compassion was 8.72. This suggests that despite experiencing the assessment to be highly compassionate overall; respecting the user, listening attentively, and treating users fairly, participants felt significantly more suspicious of the assessment, feeling their privacy was not respected, and not trusting the validity of the assessment. This may indicate an attitudinal distrust in AI systems and data privacy concerns or may be related to assessment-specific features that lead to mistrust. This finding is important as it suggests that user perception and mistrust may constitute a significant barrier to compassionate AI-powered remote care, which may lead to users and healthcare providers being less likely to adopt such technologies.

Another statistically significant finding was that those who used the chatbot during the assessment scored significantly lower on the Usability scale compared to those who did not use the chatbot feature. Participants who used the chatbot scored close to 2 whole points lower on the Usability scale (7.33) when compared to those who did not use the chatbot feature during the assessment (9.20). This may indicate that users who found the assessment confusing or less usable were more likely to ask questions to the chatbot. Another possibility is that participants found the chatbot confusing, unhelpful, or less usable and reported it on the survey, highlighting the challenges associated with support methods outside face-to-face communication. In either case, the finding suggests that the chatbot may have been unsuccessful at resolving user questions and thus increasing the overall usability of the assessment. This suggests the importance of simple, timely communication, and ease of asking questions to support user experience when designing a chatbot involved in a remote and virtual cognitive assessment. It is likely that further insights regarding chatbot experience will emerge from focus group data performed by the study group. Outside of these two significant findings, it seems that overall, each participant group experienced the assessment similarly as no significant statistical differences were observed between participant groups in any of the analyzed variables.

With respect to the third research question, these findings failed to elicit unique concerns or opportunities raised by healthcare providers with respect to AI chatbots and/or remote cognitive assessments. It is likely that the focus group portion of this study may provide more valuable insights regarding healthcare providers’ viewpoints on the rollout of such technologies.

The findings suggest a significantly reduced trust in the assessment despite higher scores on the Compassion scale, which indicates a lower level of trust overall in the assessment. These findings may be supported by Smith et al, 2016 that identified increasing the humanness of an automated agent can increase trust and usability for tasks that require social skills, such as responding to questions and providing helpful answers. The assessment experience was lacking in human characteristics such as a face or avatar that interacts with users and features a robotic voice. Increasing trust in the assessment may be attained through anthropomorphism such as the use of an interactive avatar or a more natural sounding voice. Future remote and virtual assessments may find this a beneficial way to increase trust, usability, and chatbot engagement when designing an assessment.

Another finding indicates that participants who interacted with the chatbot scored significantly lower on the Usability scale, indicating that the chatbot may have been unsuccessful at resolving questions and thus increasing the usability of the assessment. It should be noted that one participant attempted to interact with the chatbot but did not click the microphone icon and was unsuccessful at asking their question, indicating difficulty in operating and interacting with the chatbot. A systematic review of AI-chatbot use in healthcare found variability with respect to usability, acceptability, and engagement when comparing chatbot designs and protocols (Aggarwal et al., 2023) This reaffirms that ensuring that users will engage with a chatbot if they have a question and benefit from a chatbot interaction is a difficult task. Despite our chatbot being operated by a human, answers took several seconds to be produced and were read aloud in a robotic monotone voice, very unlike face-to-face human interactions. This may have been experienced negatively by users, especially during the stress of a cognitive assessment. Further research on cognitive, attitudinal, and demographic factors and their relation to chatbot experience may provide insights into human-centered universal design characteristics to support the compassionate delivery of care.

The similar overall experiences between participant or demographic groups highlight the simplicity of the CANARY experiment. Instead, the barriers seemed to be more related to remote administration, the use of Zoom, and chatbot issues. It can be noted that multiple participants had difficulty logging into Zoom and sharing their screen and audio which may have influenced their experiences. Findings on user experience scales should be taken cautiously as each scale contains a varying and low number of questions. Many of the analyzed variables did not show statistically significant results and yet many were found to have medium or large effect sizes. This is most likely due to the fact that the sample size was relatively low for this study (N=24). The variables that were found to have either a medium or large effect size indicate that practical differences may indeed exist; however, further research is required to confidently identify such patterns. Moreover, there are no consistent scales, themes, or definitions within the literature that help conceptualize user experience, so it can be difficult to compare findings to other studies.

This study is unique among the literature in evaluating the user experience and human-computer interactions with a novel, remote, AI-based cognitive assessment. Using Wizard of Oz testing and the lens of Occupational Therapy to assess user experience provides a pioneering view on remote cognitive assessment development. Providing data on user experience supports human-centered design in developing healthcare technology that is effective and caring.

The main limitations of this study are that it had a relatively small sample size (N=24) and therefore less power to its findings. Participants were selected through a sample of convenience, which led to a more highly educated group than a random sample of the population at large. Of additional note was that all participants were fluent in English and from British Columbia, Canada. This study did not collect data on participants’ ethnicity or culture. Further research may benefit from efforts to increase the enrolment of a more diverse group of individuals, such as marginalized populations, non-native English speakers, racialized minorities, and those who live in rural and remote settings. It is possible that a more diverse participant sample may provide more meaningful insights into the experience and perceptions of individuals when using AI in healthcare.

Understanding user experience, perceptions, and interactions with remote and AI-based assessments plays a critical role in operationalizing these technologies for use in healthcare. This data provides important insights into the human experience in a technologizing healthcare landscape, ensuring we can maintain core principles of accessibility, compassion, and trust through new technology adoption.

## Conclusion

This study aimed to use the Wizard of Oz method and naturalistic observations with an Occupational Therapy lens to evaluate the human-computer interactions involved in a remote and virtual AI-based cognitive assessment. The study aimed to understand how AI-assisted cognitive assessments are experienced by a variety of users and mechanisms to maintain compassionate care while incorporating technology into healthcare. Through questionnaires and naturalistic observations, the study used the Wizard of Oz method to explore the relations between the demographics, environment, experience, and perception of participants. 24 participants, 6 in each of 4 categories (patient, control, caregiver, and healthcare provider) underwent to study. Participant experience was separated into 4 main scales (Trust, Compassion, Usability, and Care Experience) and compared to self-reported demographic and data. Findings suggested that overall, members of each participant category experienced the assessment similarly and had overall positive experiences with the assessment. No significant difference was observed between survey scores and participant category, sex, device type, chatbot familiarity, and level of education. Statistically significant findings were that participants scored significantly lower on the scale Trust (8.09) than the scale Compassion (8.72) and that participants who interacted with the chatbot scored significantly lower on the Usability scales (7.33 vs. 9.20). On top of these statistically significant findings, many of the analyzed variables had medium or large effect size, indicating that participant group, participant demographics, and participant environment may play a role in user experience of this assessment. Therefore, future direction will include further exploration of user experience in self-administered remote cognitive assessments with a greater sample size.

## Data Availability

Data available upon request.

## Notes

### Competing Interest Statement

The authors have declared no competing interest.

### Funding Statement

This study was funded by AMS Healthcare.

### Author Declarations

University of British Columbia Ethics Board gave ethical approval for this work.

## References

Alzheimer Society of Canada. (2016) Report summary Prevalence and monetary costs of dementia in Canada: a report by the Alzheimer Society of Canada. Health Promotion Chronic Disease Prevention Canada. 36(10), 231–232. PMID: 27768560; PMCID: PMC5158126.

Aggarwal, A., Tam, C. C., Wu, D., Li, X., & Qiao, S. (2023). Artificial Intelligence-Based Chatbots for Promoting Health Behavioral Changes: Systematic Review. Journal of medical Internet research, 25, e40789. 10.2196/40789

Battista, P., Salvatore, C., Berlingeri, M., Cerasa, A., & Castiglioni, I. (2020). Artificial intelligence and neuropsychological measures: the case of Alzheimer’s disease. Neuroscience and Biobehavioral Reviews, 114, 211–228. 10.1016/j.neubiorev.2020.04.026

Broffman, L., Harrison, S., Zhao, M., Goldman, A., Patnaik, I., & Zhou, M. (2023). The Relationship Between Broadband Speeds, Device Type, Demographic Characteristics, and Care-Seeking Via Telehealth. Telemedicine journal and e-health: the official journal of the American Telemedicine Association, 29(3), 425–431. 10.1089/tmj.2022.0058

Chew, H. S. J., & Achananuparp, P. (2022). Perceptions and Needs of Artificial Intelligence in Health Care to Increase Adoption: Scoping Review. Journal of medical Internet research, 24(1), e32939. 10.2196/32939

Cook, S. E., Marsiske, M., & McCoy, K. J. (2009). The use of the Modified Telephone Interview for Cognitive Status (TICS-M) in the detection of amnestic mild cognitive impairment. Journal of geriatric psychiatry and neurology, 22(2), 103–109. 10.1177/0891988708328214

Di Nuovo, A., Varraso, S., Conti, D., Bamsforth J., Lucas A., Soranzo A., & McNamara J. (2019, March 11) Usability Evaluation of a Robotic System for Cognitive Testing, 14th ACM/IEEE International Conference on Human-Robot Interaction (HRI), Daegu, South Korea, pp. 588–589, doi: 10.1109/HRI.2019.8673187.

Di Nuovo, A., Varrasi, S., Lucas, A., Conti D., McNamara J., & Soranzo A. (2019) Assessment of Cognitive skills via Human-robot Interaction and Cloud Computing. Journal of Bionic Engineering 16, 526–539. 10.1007/s42235-019-0043-2

Khaligh-Razavi, S. M., Sadeghi, M., Khanbagi, M., Kalafatis, C., & Nabavi, S. M. (2020). A self-administered, artificial intelligence (AI) platform for cognitive assessment in multiple sclerosis (MS). BMC Neurology, 20(1), 193. 10.1186/s12883-020-01736-x

Kurtz, E., Zhu, Y., Driesse, T., Tran, B., Batsis, J. A., Roth, R. M., & Liang, X. (2023, June 4). Early Detection of Cognitive Decline Using Voice Assistant Commands, ICASSP 2023 - 2023 IEEE International Conference on Acoustics, Speech and Signal Processing (ICASSP), Rhodes Island, Greece, pp. 1–5, doi: 10.1109/ICASSP49357.2023.10095825.

Ihle, A., Gouveia, É. R., Gouveia, B. R., & Kliegel, M. (2017). The Cognitive Telephone Screening Instrument (COGTEL): A Brief, Reliable, and Valid Tool for Capturing Interindividual Differences in Cognitive Functioning in Epidemiological and Aging Studies. Dementia and Geriatric Cognitive Disorders Extra, 7(3), 339–345. doi: 10.1159/000479680

Minian, N., Mehra, K., Rose, J., Veldhuizen, S., Zawertailo, L., Ratto, M., Lecce, J., & Selby, P. (2023). Cocreation of a conversational agent to help patients adhere to their varenicline treatment: A study protocol. Digital health, 9, 20552076231182807. 10.1177/20552076231182807

Morrow, E., Zidaru, T., Ross, F., Mason, C., Patel, K. D., Ream, M., & Stockley, R. (2023). Artificial intelligence technologies and compassion in healthcare: A systematic scoping review. Frontiers in psychology, 13, 971044. 10.3389/fpsyg.2022.971044

Opwonya, J., Ku, B., Lee, K. H., Kim, J. I., & Kim, J. U. (2023). Eye movement changes as an indicator of mild cognitive impairment. Frontiers in Neuroscience, 17, 1171417. 10.3389/fnins.2023.1171417

Prince, M., Bryce, R., Albanese, E., Wimo, A., Ribeiro, W., & Ferri, C. P. (2013). The global prevalence of dementia: a systematic review and metaanalysis. Alzheimer’s & dementia: the journal of the Alzheimer’s Association, 9(1), 63–75. 10.1016/j.jalz.2012.11.007

Russell, S., & Novig, P. (2020). Artificial intelligence: A modern approach. Harlow: Pearson.

Sabbagh, M. N., Boada, M., Borson, S., Chilukuri, M., Dubois, B., Ingram, J., Iwata, A., Porsteinsson, A. P., Possin, K. L., Rabinovici, G. D., Vellas, B., Chao, S., Vergallo, A., & Hampel, H. (2020). Early Detection of Mild Cognitive Impairment (MCI) in Primary Care. The journal of prevention of Alzheimer’s disease, 7(3), 165–170.

Sadasivan, C., Cruz, C., Dolgoy, N., Hyde, A., Campbell, S., McNeely, M., Stroulia, E., Tandon, P. (2023). Examining patient engagement in chatbot development approaches for healthy lifestyle and mental wellness interventions: scoping review.. Journal of Participatory Medicine, 15, e45772. 10.2196/45772

Scharre, D. W., Chang, S. I., Nagaraja, H. N., Vrettos, N. E., & Bornstein, R. A. (2017). Digitally translated self-administered gerocognitive examination (esage): relationship with its validated paper version, neuropsychological evaluations, and clinical assessments. Alzheimer’s Research & Therapy, 9(1), 44. 10.1186/s13195-017-0269-3

Smith, M. A., Allaham, M. M., & Wiese, E. (2016). Trust in Automated Agents is Modulated by the Combined Influence of Agent and Task Type. Proceedings of the Human Factors and Ergonomics Society Annual Meeting, 60(1), 206–210. 10.1177/1541931213601046

Umeda-Kameyama, Y., Kameyama, M., Tanaka, T., Son, B. K., Kojima, T., Fukasawa, M., Iizuka, T., Ogawa, S., Iijima, K., & Akishita, M. (2021). Screening of Alzheimer’s disease by facial complexion using artificial intelligence. Aging, 13(2), 1765–1772. 10.18632/aging.202545

White, L., Ingraham, B., Larson, E., Fishman, P., Park, S., & Coe, N. B. (2022). Observational study of patient characteristics associated with a timely diagnosis of dementia and mild cognitive impairment without dementia. Journal of general internal medicine, 37(12), 2957–2965. 10.1007/s11606-021-07169-7

Wong, A., Nyenhuis, D., Black, S. E., Law, L. S., Lo, E. S., Kwan, P. W., Au, L., Chan, A. Y., Wong, L. K., Nasreddine, Z., & Mok, V. (2015). Montreal cognitive assessment 5-minute protocol is a brief, valid, reliable, and feasible cognitive screen for telephone administration. Stroke, 46(4), 1059–64. 10.1161/STROKEAHA.114.007253

Zygouris, S., Ntovas, K., Giakoumis, D., Votis, K., Doumpoulakis, S., Segkouli, S., Karagiannidis, C., Tzovaras, D., & Tsolaki, M. (2017). A Preliminary Study on the Feasibility of Using a Virtual Reality Cognitive Training Application for Remote Detection of Mild Cognitive Impairment. Journal of Alzheimer’s Disease: JAD, 56(2), 619–627. 10.3233/JAD-160518

